# Assessing Large Language Model Performance Related to Aging in Genetic Conditions

**DOI:** 10.1101/2025.01.19.25320798

**Authors:** Amna A. Othman, Kendall A. Flaharty, Suzanna E. Ledgister Hanchard, Ping Hu, Dat Duong, Rebekah L. Waikel, Benjamin D. Solomon

## Abstract

Unlike some health conditions that have been extensively delineated throughout the lifespan, many genetic conditions are largely described in pediatric populations, with a focus on early manifestations like congenital anomalies and developmental delay. An apparent gap exists in understanding clinical features and optimal management as patients age. Generative artificial intelligence is transforming biomedical disciplines including through the introduction of large language models (LLMs). Motivated by these advances, we explored how LLMs handle age with respect to 282 genetic conditions selected based on prevalence. We divided these conditions into five categories: Disorders limited to childhood; Disorders limited to adulthood; Disorders with changes in presentation across ages; Disorders with changes in management across ages; Disorders with no changes across ages. We evaluated Llama-2-70b-chat (70b) and GPT-3.5 (GPT) capabilities at generating accurate medical vignettes for these conditions based on Correctness, Completeness, and Conciseness as graded by 3 clinicians. Using accurately generated vignettes as in-context prompts, we further generated and evaluated patient-geneticist dialogues and assessed LLM performance in answering specific questions regarding age-based management plans for a subset of conditions. Results revealed impressive performances of 70b with in-context prompting and GPT in generating vignettes. We overall did not observe age-based biases, though our experiments identified statistically significant differences in some areas related to LLM output. Despite impressive capabilities, LLMs still have limitations in clinical applications.

## Introduction

There are currently well over 6,000 genetic conditions with identified causes, and extraordinary progress continues in identifying the specific etiologies, biological underpinnings, and manifestations of these disorders.^1^ Unlike some common health conditions, which have been comprehensively studied throughout the lifespan, many genetic conditions are typically described in pediatric populations, focusing on early and obvious manifestations like birth defects, severe biochemical or immunological perturbations, or developmental delay and other neurobehavioral findings.^2^ There are multiple reasons for this pediatric focus. One major driver is the fact that most clinical geneticists who diagnose and manage such patients are pediatrically trained.^3^ Since these clinicians primarily see, diagnose, and manage children given their scope of training, they tend to report findings in the medical literature and other outlets that pertain to the pediatric timeframe.^4^ Another explanation involves the severe nature of many genetic conditions, which frequently affects survival, especially until more recent decades, when improvements in supportive care and the availability of direct therapies for some conditions has helped enable longer lives for affected individuals.^5^ That is, many individuals with severe genetic conditions did not survive into adulthood, and thus the longer-term sequelae of these conditions were less well described. Finally, at least in some geographic areas, inequitable insurance policies mean that genetic testing is unequally covered in pediatric and adult populations, such that achieving precise diagnosis is often much more difficult in the adult population.^6,7^ As a result of these factors, gaps exist in understanding the clinical features, outcomes, and optimal management of genetic conditions as patients age.

As generative artificial intelligence (AI) continues to advance, it is rapidly transforming many biomedical disciplines, including through image generators like generative adversarial networks (GANs) for realistic image construction as well as large language models (LLMs). ^8–10^ For image generation, to produce high quality outputs that are clinically accurate, one would likely need to finetune available pre-trained models. However, many pre-trained LLMs might be used for many medical purposes without finetuning.^10–12^ Due to this versatility, the adoption of LLMs may increase accessibility to medical information across many clinical specialties by responding to queries from both medical and non-medical users.^9,13^ For example, a person with a genetic condition may ask a pre-trained LLM (e.g., GPT-4, Bard/Gemini, etc.) about future expectations or about available treatments or recurrence risks; a physician seeing a patient with a suspected genetic disorder may use these types of LLMs to better understand pertinent pathophysiology or to help generate a differential diagnosis.^14^ Going beyond single-turn conversations (a single query-and-response) with pre-trained LLMs, recent studies show that LLMs can be further developed into multi-turn chatting agents geared specifically towards medical domains, and can communicate with appropriate levels of empathy.^9,13,14^ For example, PaLM2 was finetuned on generated patient vignettes to produce a multi-turn AI chatbot with a diagnostic accuracy exceeding that of primary care physicians.^15^

Numerous studies have examined the response quality of LLMs across a spectrum of medical specialties, such as in ophthalmology, orthopedics, dermatology, obstetrics, oncology, and others. ^12,15,16^ However, none of these studies have specifically focused on genetic conditions that have age-related manifestations and management plans, and whether there can be age-related biases in the LLM responses. This is important, as LLMs may perform differently in the field of medical genomics versus other areas of medicine due to the individual rarity of genetic disorders and the relative scarcity of medical literature about many conditions.^17^ Given these considerations, we aimed to explore aspects of LLM performance in the context of medical genomics: our objective was to investigate potential age-related biases related to the identification and management of genetic conditions. To perform this study, we investigated the open-source Llama-2-70b-chat model and proprietary GPT-3.5 model in terms of proficiency of generating accurate medical vignettes encompassing 282 genetic conditions selected based on prevalence (including subgroups such as metabolic disorders). We also generated and evaluated the LLMs’ ability to generate age-appropriate patient-geneticist dialogues and their performance in answering age-specific questions regarding management with respect to the generated vignettes on a subset of these conditions. Overall, we found that LLMs perform impressively well with respect to age-related differences in our 282 conditions.

## Methods

### Dataset selection

We selected conditions based on Orphanet’s dataset of rare diseases based on prevalence (November 2023 version 2:). As shown in Figure 1, Following removal of 121 duplicates, four clinicians assessed the list of remaining conditions according to the exclusion criteria and removed those without known Mendelian/monogenic genetic causes (e.g., congenital syphilis), benign conditions that may involve a measurable phenotype, but where the clinical impact is unclear (e.g., iminoglycinuria), multifactorial conditions without a clear, known monogenic cause or where only susceptibility loci have been identified, low or unclear penetrance conditions or findings that may appear as clinical features in many different genetic conditions as well as in an isolated fashion (e.g., Hirschsprung disease), conditions caused by somatic genetic variants, such as forms of cancer (as many such conditions do not typically fall under the purview of clinical geneticists), conditions involving variable cytogenomic changes, including microdeletions or microduplications (unless clearly associated with a well-known genetic syndrome such as 22q11.2 deletion syndrome). From the initial list of 793 conditions, after applying the criteria described above, we ended with 282 conditions for analysis (See Supplementary File 1). For our analyses, we divided these 282 conditions into five categories based on information in public databases of genetic conditions including Orphanet (https://www.orpha.net/), GeneReviews (https://www.ncbi.nlm.nih.gov/books/NBK1116/), and OMIM (https://www.omim.org/), all reviewed between 02/02/2024 and 04/16/2024. These five categories are: (1) Disorders limited to childhood (age <18 years), either because most patients do not survive into adulthood (e.g., rhizomelic chondrodysplasia punctata) or the manifestations subside before adulthood (e.g., glycogen storage disease due to liver phosphorylase kinase deficiency) (n = 33); (2) Disorders limited to adulthood (age >18 years), where manifestations present later in life, although in some rare scenarios findings can appear in adolescence (e.g., GNE myopathy) (n = 13); (3) Disorders that manifest in childhood and/or adulthood, but which involve changes in presentation across the lifespan (e.g., Rubinstein-Taybi syndrome, where children have growth failure but adults have obesity)(n = 33); (4) Disorders with changes in management (with or without presentation changes) across the lifespan (e.g., homocystinuria, where adults typically require additional management considerations related to their increased risk of medical issues like strokes and osteoporosis) (n = 53); (5) Disorders with no changes in presentation or management across the lifespan (e.g., Treacher-Collins syndrome) (n = 150). For brevity, in the subsequent sections, tables, and figures, we will denote these categories as (1) Limited to childhood; (2) Limited to adulthood; (3) Presentation change; (4) Management change; (5) No change. The majority of the conditions have a neonatal onset, but many conditions have a wider age range of onset (see Figure 2 for distribution of age of onset based on data from www.orpha.net).

**Figure 1.**
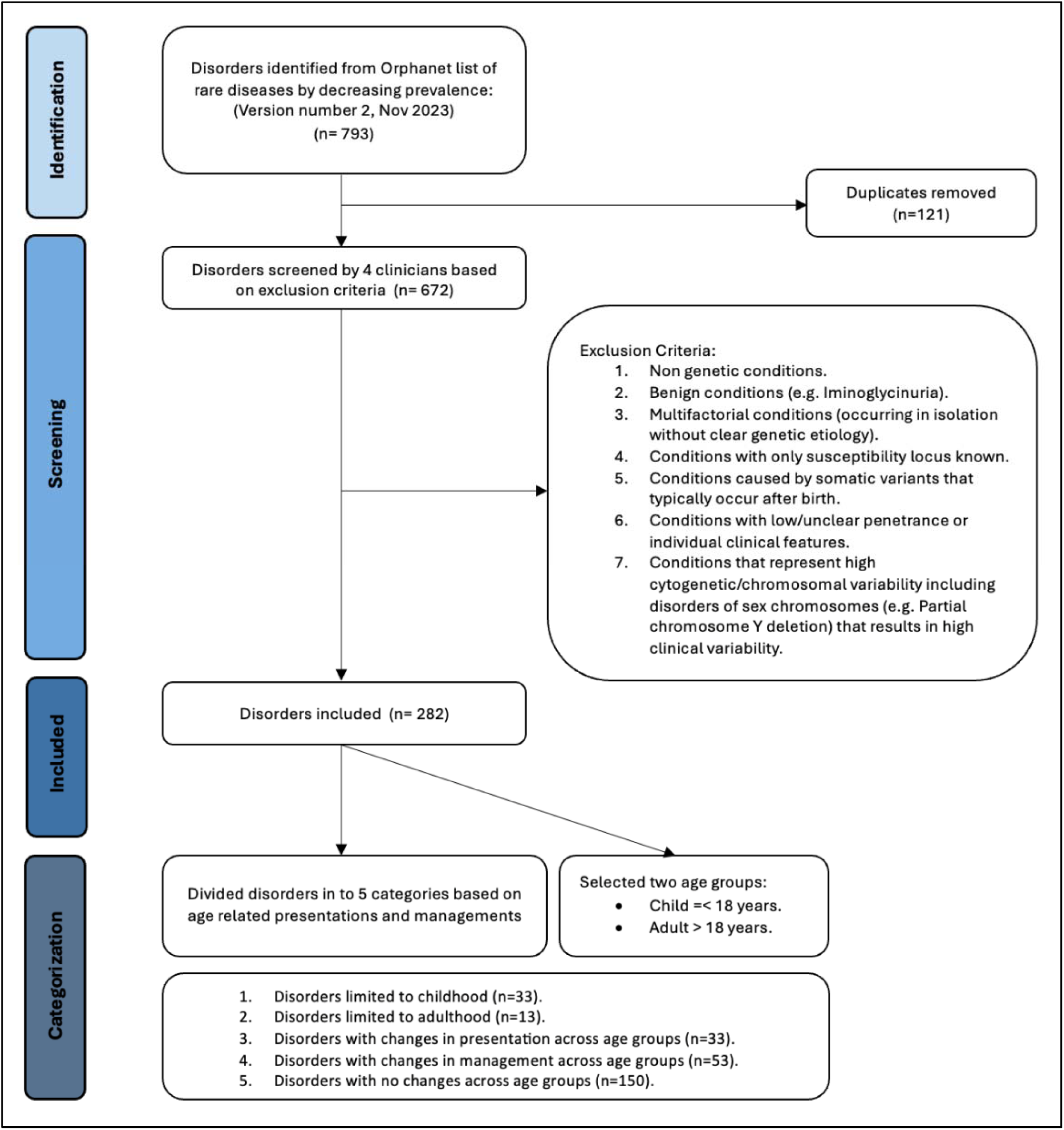
Diseases selection process, based on the PRISMA schema. This schema was adapted from the approach used for systematic reviews and is used with appropriate citation as described in the guidelines. Sources used in our analyses include Orphanet (https://www.orpha.net/), GeneReviews (https://www.ncbi.nlm.nih.gov/books/NBK1116/), and OMIM (https://www.omim.org/).

**Figure 2.**
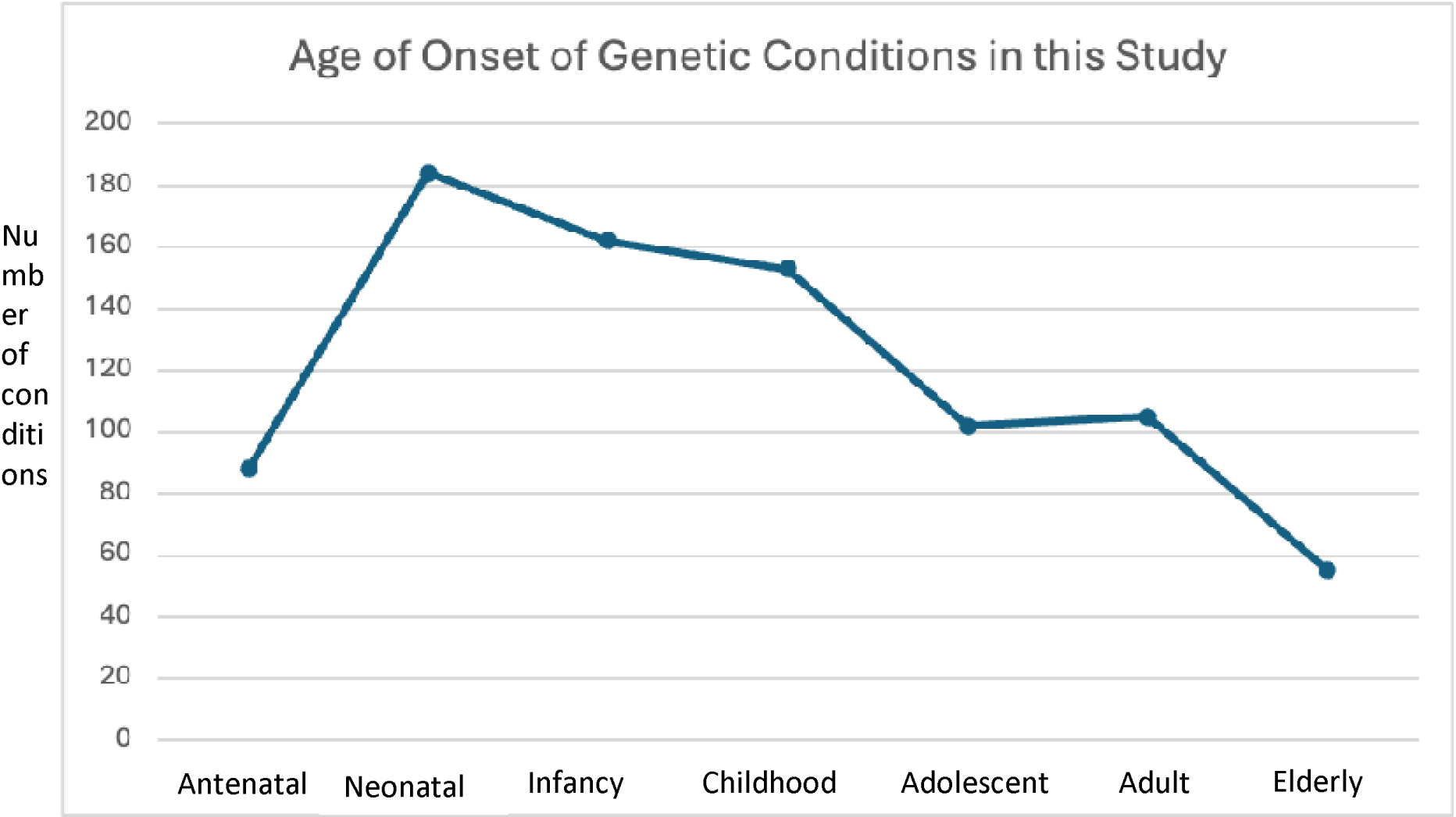
Distribution of age of onset for the 282 genetic conditions included in this study. Many conditions have a range of onset; therefore, a single condition may be represented in multiple age periods (e.g., propionic acidemia may have clinical onset during either the neonatal or infantile timeframe). Orphanet version 1.0.9 served as source for age of onset except for two cancer conditions: Gardner and Gorlin syndromes, which are now considered to be part of broader cancer disorder spectrums: familial adenomatous polyposis (FAP) and basal cell nevus syndrome (BCNS), respectively.

### Generating and scoring vignettes

We generated medical vignettes for each of our selected conditions using two LLMs: Llama-2-70b-chat and GPT-3.5. To generate vignettes with the open-source Llama-2-70b-chat, we used Biowulf, NIH’s high-performance server. Even with this server, due to the Llama-2-70b-chat model’s size, we needed to run its 8-bit C++ version to efficiently optimize memory usage and speed during data generation. The closed-source GPT-3.5 was accessed through a private Azure OpenAI instance, provided via the NIH, at a discounted institutional cost.

Two vignettes for each condition were generated by querying the LLMs: one for a patient (we use this term to refer to an individual affected by a genetic condition) during childhood and one for a patient during adulthood. First, these vignettes were generated using a “simple prompt”, without any in-context prompting. Our simple prompt was: “Generate a brief medical vignette of a [child/adult] with [genetic condition]. Incorporate the most important clinical features”, where we varied the age group of the patient (i.e., child or adult) and the name of the genetic condition.

Second, due to the cost of GPT-3.5, we applied in-context prompting only to the Llama-2-70b-chat model to generate two additional vignettes for each condition. Therefore, for each condition a total of 4 vignettes were generated by Llama-2-70b-chat (child and adult vignettes with and without in-context prompt) and 2 vignettes were generated by GPT-3.5 (child and adult vignettes without in-context prompt).

The in-context prompts for Llama-2-70b-chat were constructed using two publicly available clinical genetics databases: Orphanet (https://www.orpha.net/) and GeneReviews (https://www.ncbi.nlm.nih.gov/books/NBK1116/). We extracted information from these sources using the Orphanet Scientific Knowledge Files (https://www.orphadata.com/orphanet-scientific-knowledge-files/) and the GeneReviews NLM Open Access Subset (https://ftp.ncbi.nlm.nih.gov/pub/litarch/ca/84/), respectively. From these sources, we downloaded the relevant information for genetic conditions in XML format from both Orphanet and GeneReviews. Then, the XML files were parsed to retrieve two key descriptions: the Orphanet “Clinical description” section and the GeneReviews “Clinical characteristics” section. Some conditions in our cohort (n = 282) did not have a corresponding page in GeneReviews. In these cases, only the “Clinical description” section from Orphanet was extracted, and the GeneReviews portion was omitted. These data were then prepended with the “simple prompt” to create the full prompt with in-context data, which we will refer to as “in-context prompt”. For brevity, we will refer to the vignettes generated with in-context prompts for the child group as to as “in-context child”, and likewise “in-context adult” for the adult group (see Supplementary File 1 for simple prompts, in-context prompts, and all generated vignettes).

We note that there can be other useful resources besides Orphanet and GeneReviews. However, the Llama-2-70b-chat token limit is 4096 tokens, which constrains the total amount of in-context input data. We also did not preprocess the Orphanet “Clinical description” section and the GeneReviews “Clinical characteristics” information, for example, by removing editorial comments about specific clinical nuances, which are not fully required for the vignette generation process. Our rationale is that the average user would likely use the in-context data as-is. Moreover, having more related information in the in-context prompt typically improves the LLM output.^18,19^

Previous studies have introduced approaches for using an LLM to grade its own output (or output from another LLM).^15,20^ For example, one can auto-evaluate the generated vignettes by using in-context learning with few-shot examples (e.g., 5 in-context examples).^15^ In-context learning ideally requires accurate human manual grading for several vignette instances of the same disease; moreover, these instances should contain both good and bad vignettes.^21^ However, for our approaches, as it can require significant amount of time and expertise to grade a single vignette, and as we aimed to evaluate LLMs on many conditions under various settings (e.g., pediatric versus adult, with and without in-context prompting), we decided to focus on grading a single vignette from many different diseases, rather than grading many different vignettes from the same disease.

Second, in-practice, for the same prompt, we did not observe a wide range of variability among the vignettes when using different random seeds with Llama-2-70b-chat (See Supplementary Table 1 for the results of repeated runs). Roberta vector embedding was used to compare the first generated vignette against each subsequent repeated runs with different random seeds.^22^ There can be other kinds of document embedding; however, Roberta was used successfully in similar studies.^23^ Supplementary Table 1 shows high similar scores among the vignettes generated with the same prompt but with different random seeds.

This low variability implies that, for future studies, we would need to carefully design different in-context prompts for the same disease so that the corresponding vignettes would be at varying levels of accuracy. Ideally, in-context prompts should not be fully unrelated to the disease descriptions; thus, we need to carefully determine which plausible phenotypes to include or exclude from the in-context prompt. This is a difficult task because the accuracy can be equally affected not only by omitting rare phenotypes (manifestations) but also by omitting common phenotypes. For example, Wilson disease contains the rare phenotype “Kayser-Fleischer ring” which might be considered a “pathognomonic” feature (i.e., a feature that greatly helps in identifying a specific disease). Conversely, Rett syndrome contains the common phenotypes: developmental delay, mobility issues, and repetitive hand movements. Thus, with Rett syndrome, it is not immediately obvious which phenotype or set of phenotypes should be included or excluded. Moreover, the grading criteria is also complicated by the fact that some conditions have many more associated phenotypes than others.

Third, most open-source models do not have a large token limit, and thus cannot handle many vignette examples for in-context learning. Proprietary models with large token limit would require significant additional computation cost. For these reasons, we manually graded each vignette, and reserve the topic of auto-evaluation for future studies.

Manual scoring of the vignettes was performed by 3 clinicians; each graded 6 different vignettes for approximately one-third of the 282 conditions; child (70b), adult (70b), child (70b context), adult (70b context), child (GPT), and adult (GPT) vignette. A scoring rubric was used to grade the vignettes to help ensure consistency. There are multiple ways to score LLM output; we used a version of a method previously described (https://docs.rungalileo.io/galileo/gen-ai-studio-products/galileo-evaluate), as we felt that it adequately provides a way to judge an LLM output. We assessed each vignette for Correctness, Completeness, and Conciseness, assigning a score of 0 (not correct, complete, or concise) or 1 (correct or complete or concise), yielding a maximum total score of 3 for each. Since all results received 1 scores for conciseness, we removed this consideration from scoring the vignettes (See Supplementary Table 2 for vignette scoring rubric). Correctness score measures whether a given response is factual. That is, we want to avoid “hallucinations”, which are presentations of information that are logically consistent and could easily be believable to a nonexpert, but which are factually incorrect (e.g., a vignette that describes an individual affected with campomelic dysplasia to have long limbs, whereas campomelic dysplasia is actually associated with short limbs). Completeness score measures how thoroughly the LLM response covered important features relevant to the genetic disorder. Considering that not all individuals affected by a genetic condition display all the textbook characteristics, but instead demonstrate varying degrees of severity and different manifestations, clinicians evaluating the vignettes considered a vignette complete if it exhibited the most essential features (based on the sources we used) along with sufficient additional clinical and laboratory indicators for a given condition. For example, a vignette of a patient with Freidreich ataxia would be considered incomplete if the description does not include impaired muscle coordination (ataxia) and impaired speech (dysarthria). To enable additional scoring, a separate metric “accuracy” was marked as 1 if a vignette receives 1 for both correctness and completeness.

To ensure consistency and agreement between the three clinicians who graded the vignettes, independent scoring of ten conditions were done by a combination of each two clinicians (Clinician 1&2, Clinician 1&3, Clinician 2&3). Kappa statistic was used to measure inter-rater reliability which showed that all 3 clinicians did not have significant disagreement^24^ (See Supplementary Table 3).

To assess for the presence of other biases within the LLM generated vignettes, we extracted identifiers from within the vignettes (namely: age and gender) and compared them to each condition’s characteristic age of onset, sex-specific prevalence, and mode of inheritance using Orphanet database (https://www.orpha.net/en/disease) as the source. The presence of this type of genbias was identified when the isolated vignettes’ variables did not match the conditions’ specific characteristics. Gender rather than biological sex was extracted from vignettes because less than 1% presented biological sex identifiers (e.g., 46, XY), whereas 96.8% of vignettes did provide gender identifiers (e.g., his and her). Given gender and biological sex are roughly 50% male and 50% female in the general population, deviations from this ratio in vignette gender output was considered gender bias. Conditions that preferentially affect one sex were also considered when examining gender bias.

### Generating and scoring dialogues

Using Llama-2-70b-chat, we designed a self-conversation environment to simulate a medical dialogue between a patient/family and a geneticist with the generated vignettes as input background information. Dialogues were only generated for disorders in two categories: presentation change and management change. Furthermore, within these categories, dialogues were only generated for conditions whose vignettes scored a 1 for both correctness and completeness (i.e., accurate vignettes) when graded by our clinicians. This is because these vignettes are used as the initial prompt to provide background information to the LLM when generating the dialogues. In other words, the patient in each generated vignette becomes the patient visiting a geneticist in our dialogues, and the LLM builds the patient-geneticist conversation off the background information from this vignette. A total of 41 disorders fit these criteria and had dialogues generated for them.

The same Llama-2-70b-chat instance plays both the role of the patient/family (Agent 1) and the geneticist (Agent 2), in a multi-turn conversation with itself. Through the Llama-2-70b-chat infrastructure, the ‘system’ input argument, which is not part of the prompt, provided the LLM instructions for both roles. Figure 3 illustrates how these instructions were given to each chat agent.

**Figure 3.**
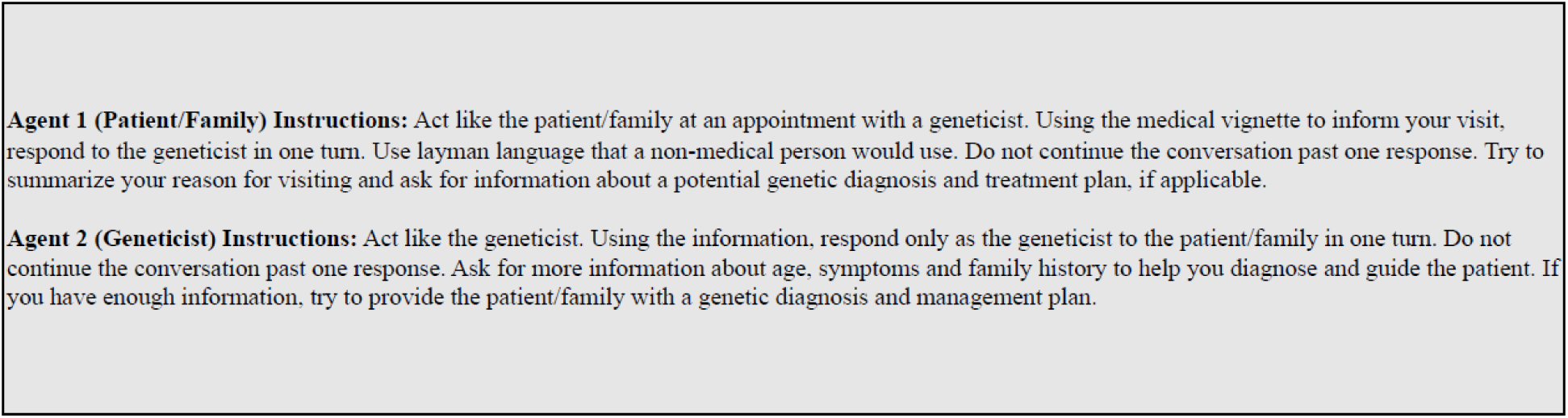
Instructions given to each agent in the self-conversation process to generate a dialogue using Llama-2-70b-chat, based on an accurate medical vignette. Instructions were presented to each instance of Llama-2-70b-chat, for a maximum of 6 turns to produce each dialogue.

With the generated medical vignettes as prompts, the patient/family agent describes the clinical manifestations and reasons for visiting the geneticist and asks for any relevant medical questions. The geneticist agent would then offer guidance towards potential differential diagnoses, and management options. The geneticist agent was specifically instructed to ask for pertinent questions to the patient/family agent to reveal more relevant medical information.

Figure 4 summarizes this multi-turn conversation procedure with an example of a child vignette about the condition Stickler syndrome. The dialogue starts with Llama-2-70b-chat acting as the patient/family agent initiating the conversation, by using the vignette as in-context prompt to summarize their clinical features and ask for guidance. Llama-2-70b-chat then acts as the geneticist agent, and its corresponding reply is appended into the dialogue history. Due to token limit, the conversation continues for a maximum of 6 turns, allowing 3 responses per agent. All previous outputs are appended to the dialogue history and used as in-context information for the next output. However, the patient vignette is only visible to the LLM when the patient/family agent is responding, as referential information about the condition. Throughout the dialogue, the geneticist role does not have access to the patient vignette; it only has access to the information provided by the patient/family agent.

**Figure 4.**
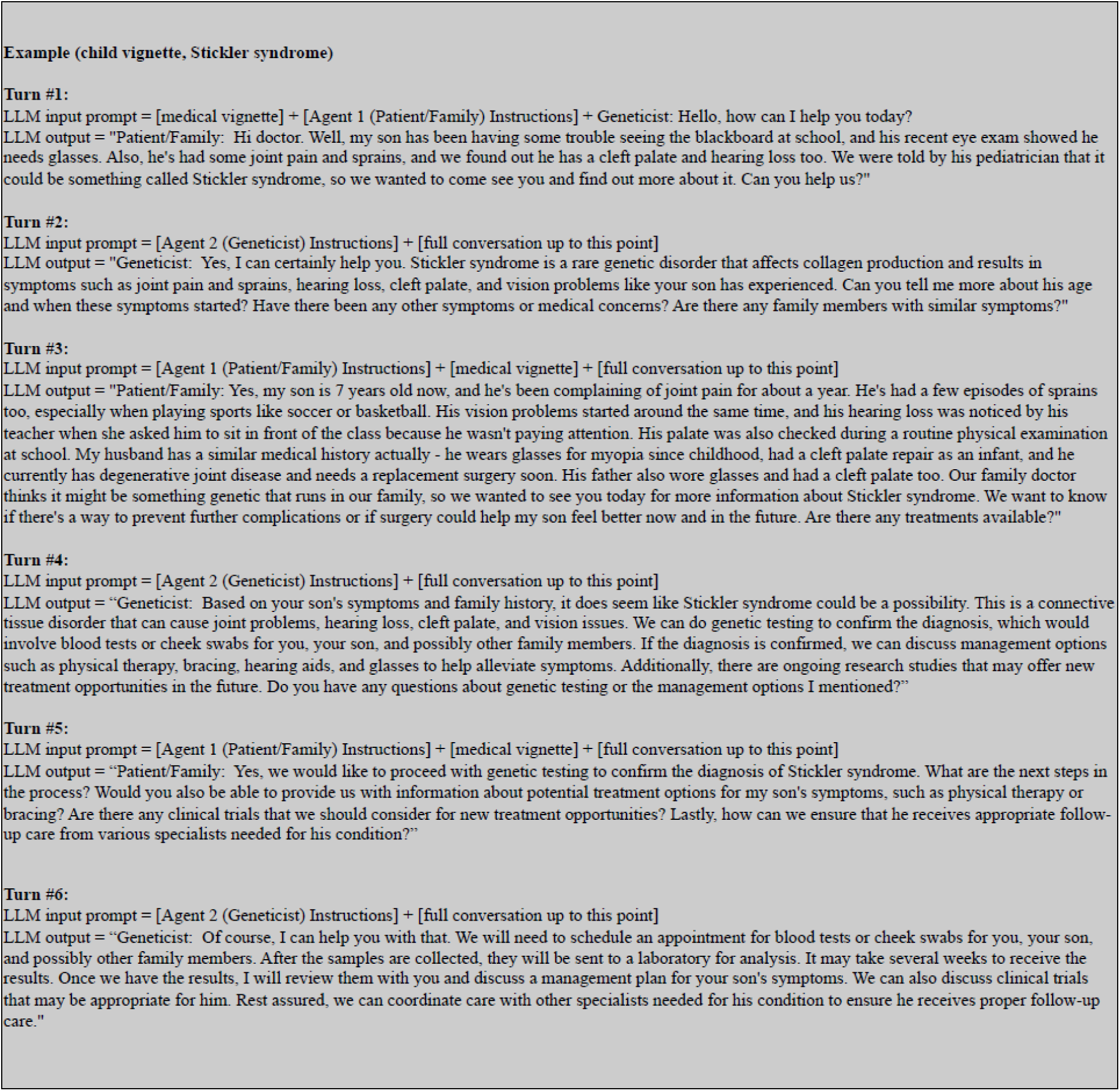
Llama-2-70b-chat example self-conversation process to generate a dialogue between the patient/family and geneticist, based on an accurate medical vignette. See Supplementary File 1 for full dialogues and vignettes. The bracket indicates a shortened representation of the input; for example, [medical vignette] represents the entire generated vignette of a child with Sticker syndrome. For each instance of the geneticist agent, the [full conversation up to this point] does not contain the [medical vignette].

Considering the auto-evaluation difficulties confronted when scoring vignettes, grading of the generated dialogues was also done manually and by only one clinician since the inter-rater reliability showed no significant disagreement based on Kappa statistic performed on the vignettes scoring (see Supplementary Table 3). Because dialogues were longer and more detailed than the vignettes, we applied a scoring rubric to assess three metrics; 1) Correctness, which assessed how realistic and appropriate the clinical description and management recommendations relevant to the genetic diagnosis, patient age and gender; 2) Completeness, which assessed if the dialogue provides a complete list of the important clinical, laboratory features and management options relevant to the genetic disorder; 3) Compassion, which assessed if the conversation shows appropriate empathy and reassurance, provides resources and avoids the use of medical jargons or other terminology that would be very challenging for a layperson to understand. Each of the three metrics was graded on a 5-point Likert scale (ranging from 1 for strongly disagree to 5 for strongly agree) to match and capture the overall length of the dialogue. (See Supplementary Table 4 for dialogue scoring rubric)

### Generating and Scoring Management Plans

Considering the easy accessibility of LLMs to the public, including their use by patients as well as the wide reliance of physicians on online resources to aid in medical decision making and guide patient management^11,12,16,25^, we decided to specifically test LLM ability in answering direct questions about management recommendations for genetic conditions while considering different age groups. For this purpose, we selected conditions that fell into the category of “management change” (n=53) and generated two responses for each of these conditions, one for a child and one for an adult. We prompted Llama-2-70b-chat to provide a management plan for a patient with each condition. This hypothetical patient was assigned the same age and gender from the generated vignette. We note that we did not provide the full generated vignettes or any other data as in-context prompt. We simply asked the LLM to provide appropriate recommendations including medications, genetic testing, and specialist referrals. This was done to simulate a more realistic usage of an LLM from the average user. It is unlikely that a patient or family member of someone with a genetic condition would source and use highly specialized in-context prompting when using an LLM.

These management plans were manually scored by one clinician using the same rubric used for scoring the vignettes (See Supplementary Table 2), with Correctness, Completeness, and Conciseness all considered, for a maximum of 3 total points. For conditions with management plans that received a low score (0 or 1 out of 3 total points) on Llama-2-70b-chat, we used GPT-3.5 as another option to generate alternative answers. The goal is to observe whether GPT-3.5 would provide more accurate results for the cases in which Llama-2-70b-chat performs poorly.

### Data availability

All data are available in the main manuscript, accompanying figures, tables and supplementary materials. Code to download the model, generate Llama-2-70b-chat responses (i.e., vignettes, dialogues, management plans), and run statistical tests can be accessed at: https://github.com/flahartyka/LLM-aging-in-genetic-conditions

## Results

### Evaluating generated vignettes

In this section and onward, for brevity, we will denote GPT-3.5 as GPT, and the experiment settings Llama-2-70b-chat with and without in-context prompts as 70b and 70b Context, respectively.

The Correctness, Completeness, Total score, and Accuracy of the vignettes generated by 70b, 70b Context and GPT were tallied using the described clinical grading rubric (Supplementary Table 2). Correctness and Completeness combine to form the Total score, while Accuracy is a measure of total vignette success—the Accuracy score is only counted as 1 if both the Correctness and Completeness score receive a 1. To compare between two outcomes, we conducted t-test (either paired or unpaired t-test depending on the groups being compared) and applied Bonferroni correction on the standard 5% false positive threshold.

We first evaluated whether age-bias exists (i.e., comparing child to adult group) for each LLM setting with respect to each of the five disease categories “Limited to Childhood”, “Limited to Adulthood”, “Presentation Change”, “Management Change”, and “No Change” (Figures 5 and 6).

**Figure 5.**
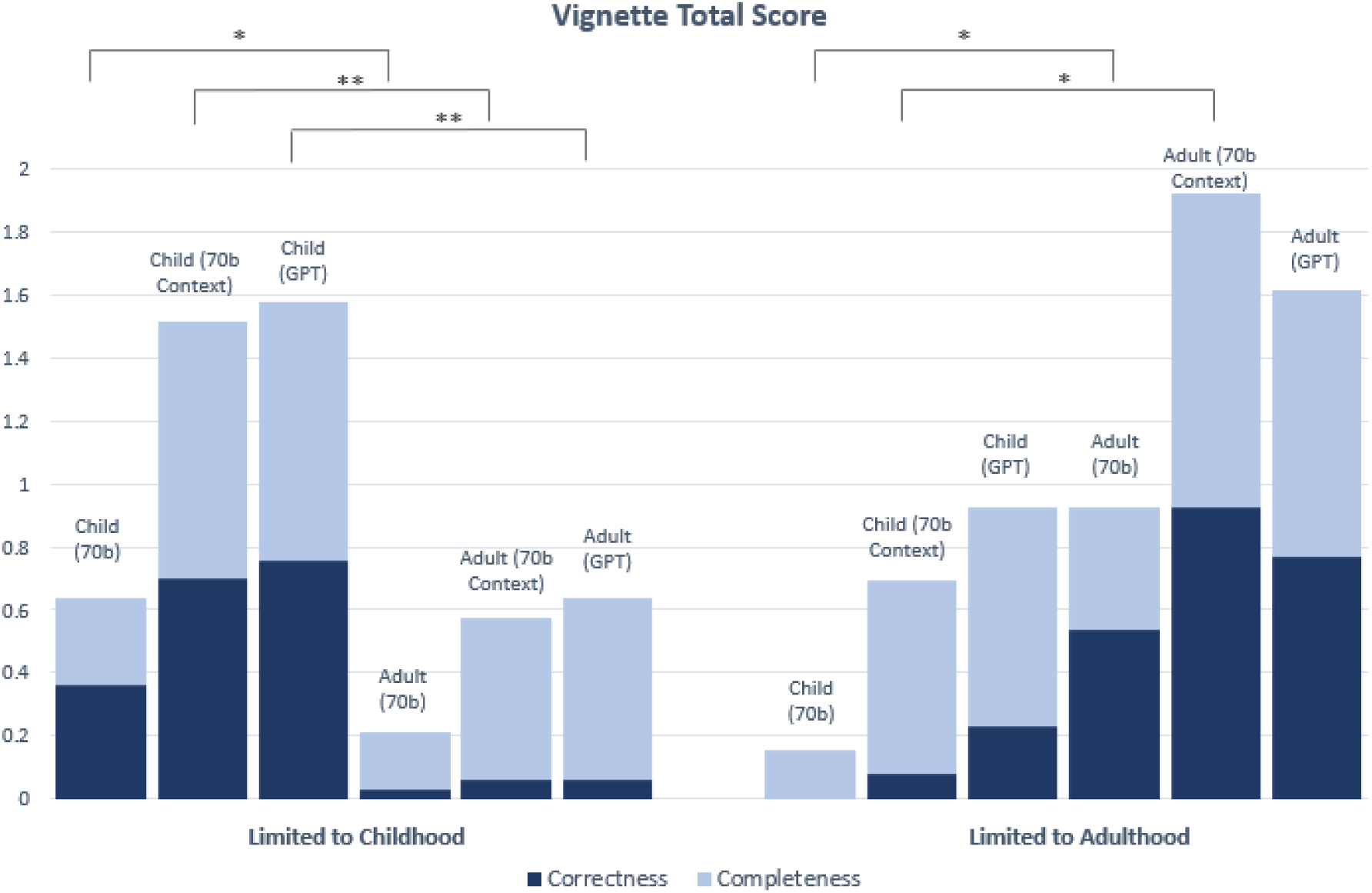
Correctness and Completeness score breakdowns for vignettes in “Limited to Childhood” and “Limited to Adulthood” categories. All vignettes were clinician-graded and assigned a score of either 0 or 1 for Correctness and Completeness, for a total of 2 possible points. Significance (*) is indicated at the α = 0.05 level between child and adult vignettes for the Correctness score only. Two markers (**) indicates significance at the α = 0.05 level between child and adult scores for both the Correctness and Completeness. In this chart, significance is only demonstrated for comparisons between child and adult vignettes for the same model (See Supplementary Tables 5 and 6 for statistical comparisons between 70b, 70b Context and GPT). The Bonferroni threshold for significance is p < 0.005. This chart shows the combined Correctness and Completeness scores; see Supplementary Figures 1 and 2 for individual Correctness and Completeness graphs with variance.

**Figure 6.**
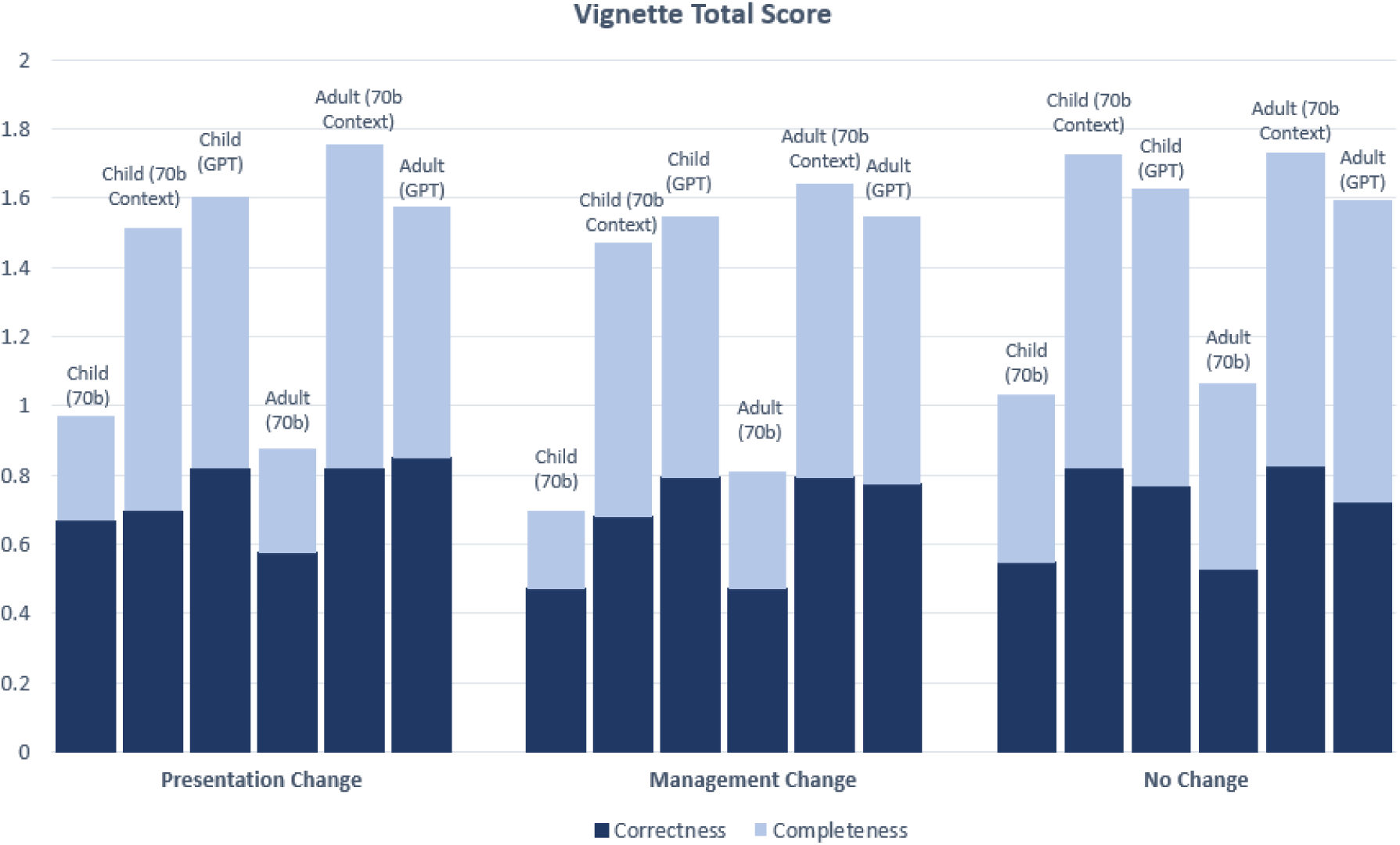
Correctness and Completeness score breakdowns for vignettes in “Presentation Change”, “Management Change”, and “No Change” categories. All vignettes were clinician-graded and assigned a score of either 0 or 1 for Correctness and Completeness, for a total of 2 possible points. Significance (*) is indicated at the α = 0.05 level between child and adult vignettes for the Correctness score only. Two markers (**) indicates significance at the α = 0.05 level between child and adult scores for both the Correctness and Completeness scores. In this chart, significance is only demonstrated for comparisons between child and adult vignettes for the same model (See Supplementary Tables 5 and 6 for statistical comparisons between 70b, 70b Context and GPT). The Bonferroni threshold for significance is p < 0.005. This chart shows the combined Correctness and Completeness scores; See Supplementary Figures 1 and 2 for individual Correctness and Completeness graphs with variance.

With regard to the Correctness score, when conditioned on the same LLM setting (70b, 70b Context, or GPT), averaging over the diseases “Limited to Childhood”, the generated child vignettes had statsistically higher Correctness scores than the adult vignettes (p= 0.00123, p=1.61 × 10^-8^, p=8.37 × 10^-10^ in 70b, 70b Context and GPT, respectively). This is expected because LLMs are not likely to generate reliable descriptions of adult patients for such diseases. The expected and opposite trend was observed for diseases “Limited to Adulthood”, where vignettes for adults on average obtained higher Correctness score than vignettes for the child group. Two of three comparisons between child and adult vignettes in “Limited to Adulthood” are statistically significant (p= 0.00281, p=3.21 × 10^-6^, p= 0.0124 in 70b, 70b Context and GPT, respectively) (See Figure 5. See Supplementary Figure 1 for individual Correctness graphs).

Specifically, for conditions in the category “Limited to Childhood” and “Limited to Adulthood”, the LLMs generated a description of a person’s age that does not correlate properly with the associated condition category and hence was graded 0 for Correctness (e.g., the LLM models generated a vignette describing a 10-year-old child vignette for amyotrophic lateral sclerosis (ALS), a progressive neurodegenerative disease that typically affects adults, and a vignette describing a 35-year old adult for rhizomelic chondrodysplasia punctata, a condition in which it is rare for an affected individual to live past age 10).

In terms of Correctness score for conditions in the other three categories “Presentation Change”, “Management Change”, and “No Change”, there was overall high performance, particularly for 70b Context and GPT generated vignettes (with Correctness score ranging between 68%-85% for these two models), with no statistically significant differences between the child and adult vignettes’ Correctness scores among all three types of experiments (70b, 70b Context, GPT) (See Figure 6. See Supplementary Figure 1 for individual Correctness graphs).

Considering the Completeness score, there were statistically significant differences between some of the child and adult vignettes for the “Limited to Childhood” category only. Two of the three comparisons between child and adult vignettes in “Limited to Childhood” are statistically significant (p=0.000743 for 70b Context, and p=0.00915 for GPT). There was no significant difference in Completeness score between the child and adult vignettes for the “Limited to Adulthood”, “Management Change”, “Presentation Change”, and “No Change” disease categories with overall high performance, particularly for 70b Context and GPT generated vignettes (with Completeness score ranging between 73%-94% for those two models) (See Figure 5. See Supplementary Figure 2 for individual Completeness graphs).

The Accuracy score was only counted as 1 if both the Correctness and Completeness score received a 1. Statistically significant differences were observed between the Accuracy of child and adult vignettes for two of three comparisons (70b Context, GPT) made in the “Limited to Childhood” category (p=1.99 × 10^-7^ for 70b Context, and p= 3.93 × 10^-9^ for GPT). Statistically significant differences were observed between child and adult vignettes for one of three comparisons (70b context) made in the “Limited to Adulthood” category (p=3.21E-06 for 70b Context) (Figure 7). There was no statistically significant difference between the Accuracy of child and adult vignettes for the disease category “Presentation Change”, “Management Change”, or “No Change” (Figure 8).

**Figure 7.**
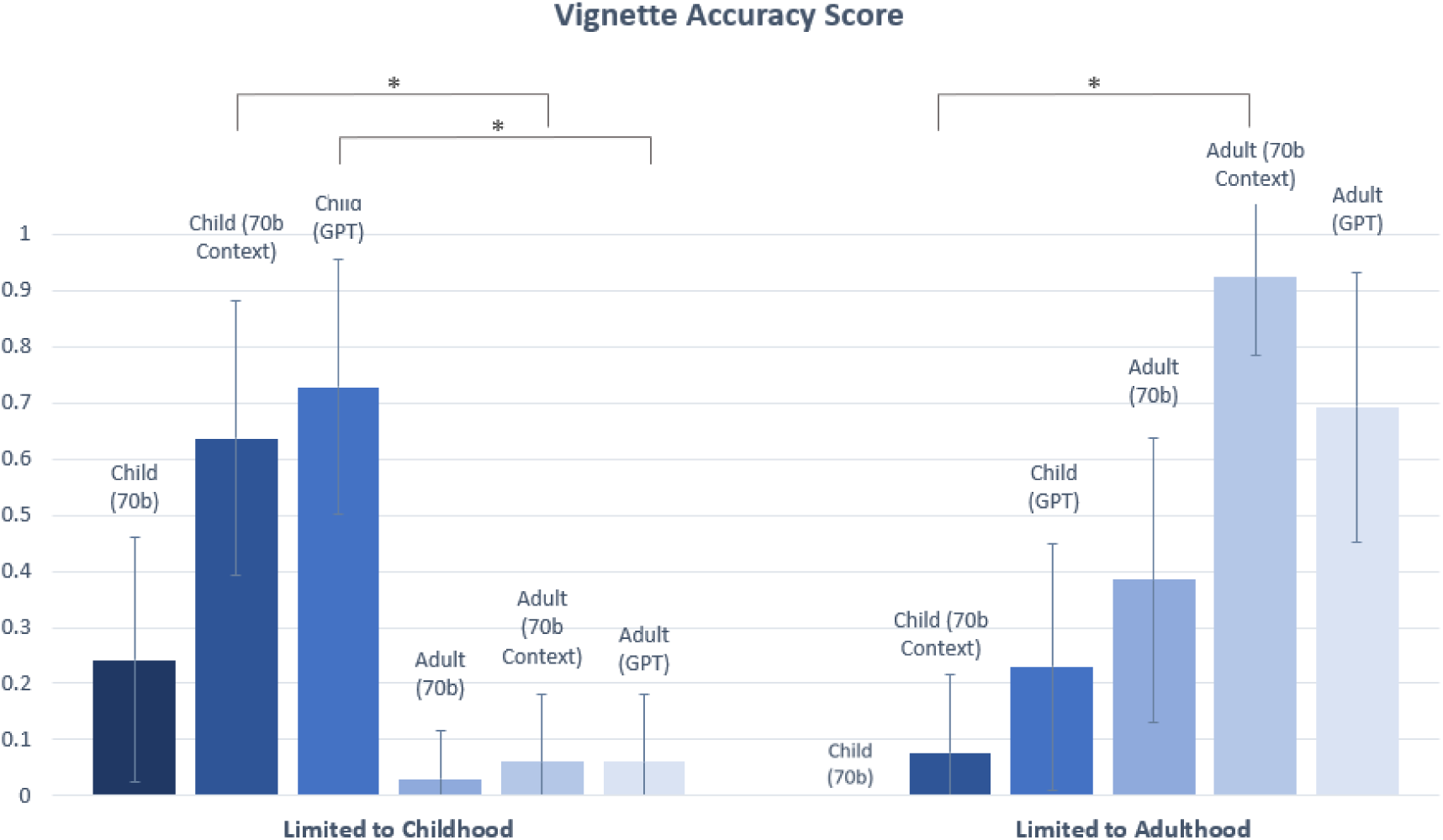
Accuracy scores for vignettes in “Limited to Childhood” and “Limited to Adulthood” categories. Statistically significant differences (*) in Accuracy scores are indicated at the α = 0.05 level between child and adult vignettes. In this chart, significant differences are only demonstrated for comparisons between child and adult vignettes for the same model (See Supplementary Tables 5 and 6 for statistical comparisons between 70b, 70b Context and GPT). The Bonferroni threshold for significance is p < 0.005.

**Figure 8.**
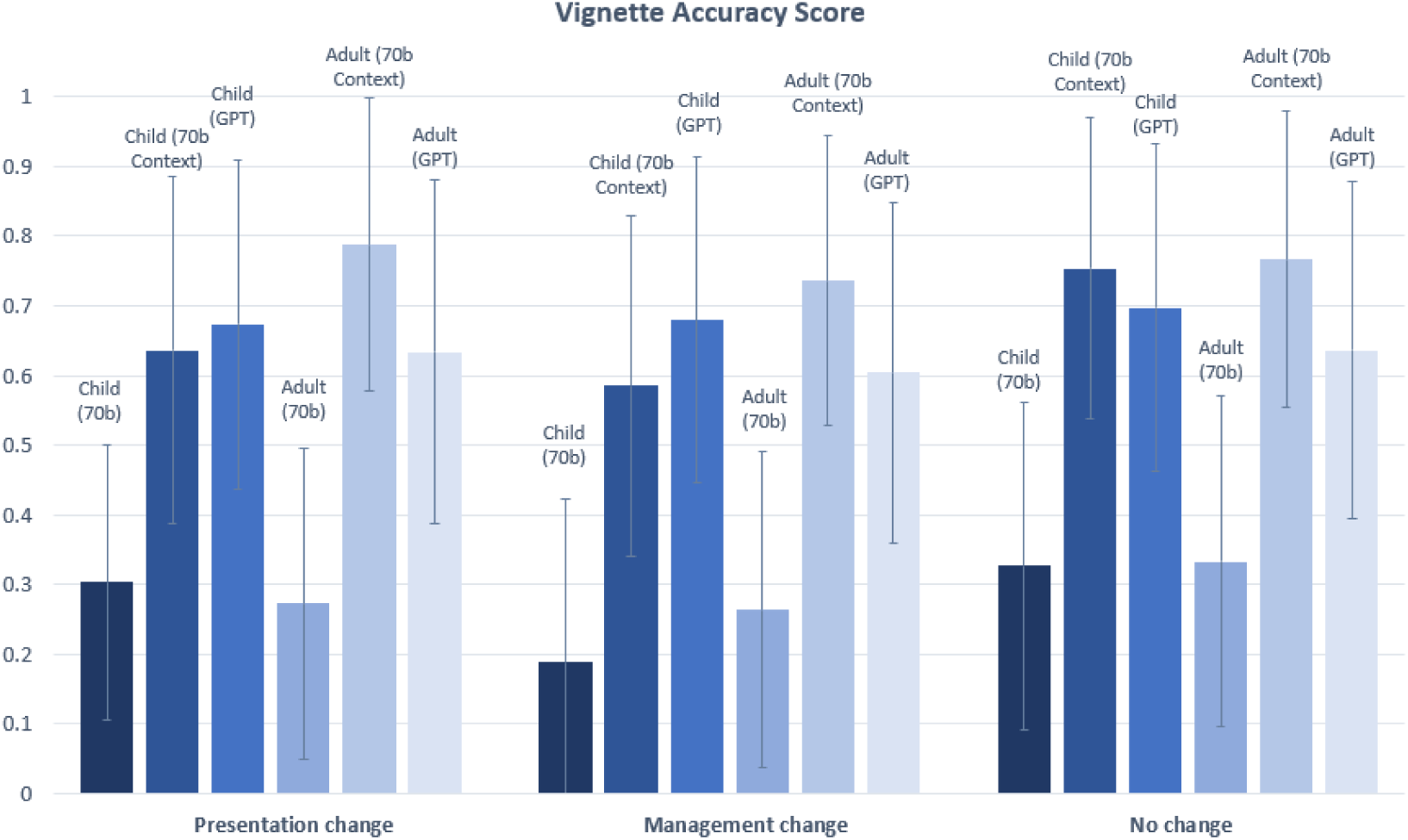
Vignette Accuracy score for vignettes in “Presentation Change”, “Management Change”, and “No Change” categories. Statistically significant differences (*) in Accuracy scores are indicated at the α = 0.05 level between child and adult vignettes. In this chart, significant differences are only observed for comparisons between child and adult vignettes for the same model (see Supplementary Tables 5 and 6 for statistical comparisons between 70b, 70b Context and GPT). The Bonferroni threshold for significance is p < 0.005.

Overall, 70b, 70b Context and GPT comparisons between child and adult vignettes exceeded our initial expectations and did not show significant age-bias in generating descriptions of genetic conditions.

When comparing the three types of experiments (70b, 70b Context, GPT), the general trend shows that leveraging in-context prompts improves Llama-2-70b-chat, especially for the Completeness score metric. For child vignettes, 70b Context scored significantly higher than 70b in Completeness in four of five total disease categories (p=1.58 × 10^-5^ for the “Limited to Childhood” category, p=1.02 × 10^-5^ for the “Presentation Change” category, p=4.28 × 10^-10^ for the “Management change” category, and p=8.66 × 10^-17^ for the “No Change” category).

Similarly, when evaluating the adult vignettes, 70b Context also scores significantly higher than 70b in Completeness in four of five total disease categories (p=0.000893 for the “Limited to Adulthood” category, p=1.61 × 10^-8^ for the “Presentation Change” category, p=3.86 × 10^-8^ for the “Management Change” category, and p=7.76 × 10^-13^ for the “No Change” category) (See Supplementary Table 5). This implies that Llama-2-70b-chat may not have been fully trained specifically on the data related to these criteria; hence, without in-context prompting, it does not perform consistently well. GPT-3.5, however, obtains very high Accuracy scores without any prompting. While the Llama-2-70b-chat in-context prompting accuracies have higher average scores than GPT-3.5, these differences are non-significant (Supplementary Table 6). This implies that (1) in-practice, although Llama-2-70b-chat is free and open-source, it may be easier to use GPT-3.5 to avoid finding in-context prompting information and (2) with proper prompting, Llama-2-70b-chat performance can rival and outperform GPT-3.5.

### Mean Age presented in vignettes for the “Limited to Childhood” and “Limited to Adulthood” categories

We performed a sub-analysis to examine mean age for the described individuals in the child and adult vignettes generated by the LLMs for “Limited to Childhood” and “Limited to Adulthood” vignettes (See Figure 9). Comparing 70b to 70b Context for child vignettes in conditions “Limited to Childhood”, the mean age decreased from 3.2 years to 1.4 years when in-context prompting was added, which was a statistically significant difference (p=2.32 × 10^-5^).

**Figure 9.**
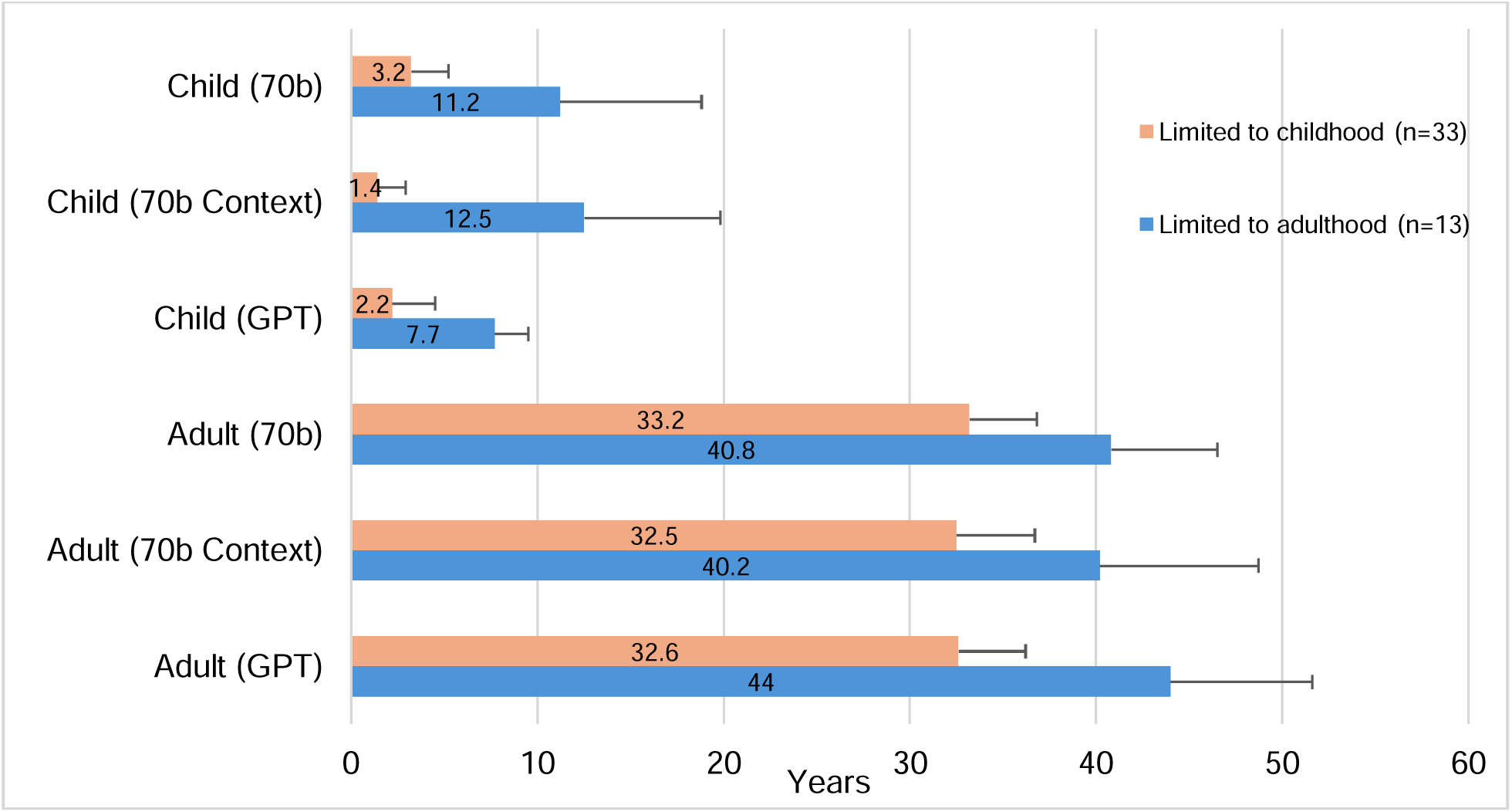
Mean age in years presented in vignette output was calculated for “Limited to Childhood” and “Limited to Adulthood” conditions. “Limited to Childhood” condition is defined as either limited survival due to disease severity or as manifestations may resolve prior to the second decade of life. A “Limited to Adulthood” condition is defined as a condition that presents in adolescence or later. Standard deviation of age is presented in the error bar only shown in the positive direction. Significance (*) is indicated at the α = 0.05 level, between 70b, 70b Context, and GPT for each age group. The Bonferroni threshold for significance is p < 0.00833.

Additionally, the mean age decreased from 3.2 years to 2.2 years when comparing child vignettes generated through 70b and GPT, respectively; this was also statistically significant (p=0.005401). For adult vignettes in the “Limited to Adulthood” category, significant differences were found between one comparison of mean ages: 70b Context vs GPT. The mean age for adult vignettes is 40.8 for 70b. When context is added, there is a nonsignificant change, as the mean age is decreased to 40.2. Using GPT, the mean age for adult vignettes is 44, which is significantly higher than 70b context (p=0.00311), but not significantly higher than 70b (p=0.0863).

For conditions in the “Limited to Adulthood” category, 70b, 70b Context, and GPT inaccurately generated vignettes for patients with a child age (< 18 years) when asked for child vignettes, even though the conditions in this category only manifest in adults. However, they all generated vignettes with relatively older child ages (mean age: 7.7-11.2 years), when compared to child vignettes within the “Limited to Childhood” category (mean age: 1.4-3.2 years). This reflects that the LLMs have likely been trained that these disorders tend to present later in life. Similarly, in the “Limited to Childhood” category, adult vignettes, with patients aged >18 years old, were inaccurately generated for conditions in this category, where most patients either die in childhood or the manifestations resolve prior to adulthood. These incorrect vignettes were generated across all three experiments (70b, 70b Context, GPT); however, the patients in these generated vignettes had a relatively younger adult age (mean age: 32.5-33.2 years), when compared to the adult vignettes for the “Limited to Adulthood” category (mean age: 40.2-44 years). This again reflects that LLMs are at least partially trained on data that indicates that these conditions are most likely to present at younger ages.

In two instances, GPT appropriately refused to provide these contradicting vignettes. It provided the following response when asked to generate a child vignette for a condition from the “Limited to Adulthood” category “ALS: I’m sorry, but it’s not possible for a child to have Amyotrophic lateral sclerosis (ALS). ALS is a progressive neurodegenerative disease that typically affects adults, and its onset is rare in individuals younger than 20 years of age…”. It also provided the following response when asked to generate an adult vignette for a condition from the “Limited to Childhood” category, “Infantile myofibromatosis is a rare condition that typically affects infants and young children. However, in rare cases, it can also affect adults. Here is a brief medical vignette of an adult with infantile myofibromatosis…”.

### Mean age presented in child vignettes for metabolic conditions

From the original cohort of 282 conditions (disregarding the 5 disease categories), we evaluated LLM performance on the child vignettes conditioned only on inherited metabolic diseases (sometimes called “inborn errors of metabolism” or “biochemical disorders”), which are genetic conditions that result from a missing or defective enzyme in the body, disrupting how the body makes or uses proteins, fats, or carbohydrates. These conditions typically present very in early life. They are also important medically because several of these conditions can lead to death at a very young age. We analyzed the mean age presented in the child vignettes for three groups of metabolic diseases (1) “All metabolic conditions” contained within our original cohort, n=74; (2) a sub-group of “metabolic conditions listed within the Recommended Uniform Screening Panel (RUSP) for newborn screening (NBS)” as of April 22, 2024 (e.g., Phenylketonuria), n=21 (https://www.hrsa.gov/advisory-committees/heritable-disorders/rusp); (3) a smaller sub-group of “metabolic conditions with acute neonatal crisis” where these acute events are very important medical issue that tend to present soon after birth (e.g., Maple syrup urine disease (MSUD), n=11 (See Supplementary File 1). We intentionally focused on analyzing child vignettes rather than adult vignettes as most of these metabolic conditions have childhood onset. The results showed a statistically significant decrease in mean age generated by 70b Context from 3.9 years to 0.8 years between “all metabolic conditions” group versus the group of “metabolic conditions with acute neonatal crisis” (p= 0.006481). Additionally, there was a similar statistically significant decrease in mean age generated by GPT from 3.9 years to 1.2 years between the two groups (p= 0.002258). We also observed a statistically significant decrease of mean age for the “metabolic conditions with acute neonatal crisis” (from 2.1 years to 0.8 years) when comparing 70b to 70b context (p= 0.00565) (See Figure 10).

**Figure 10.**
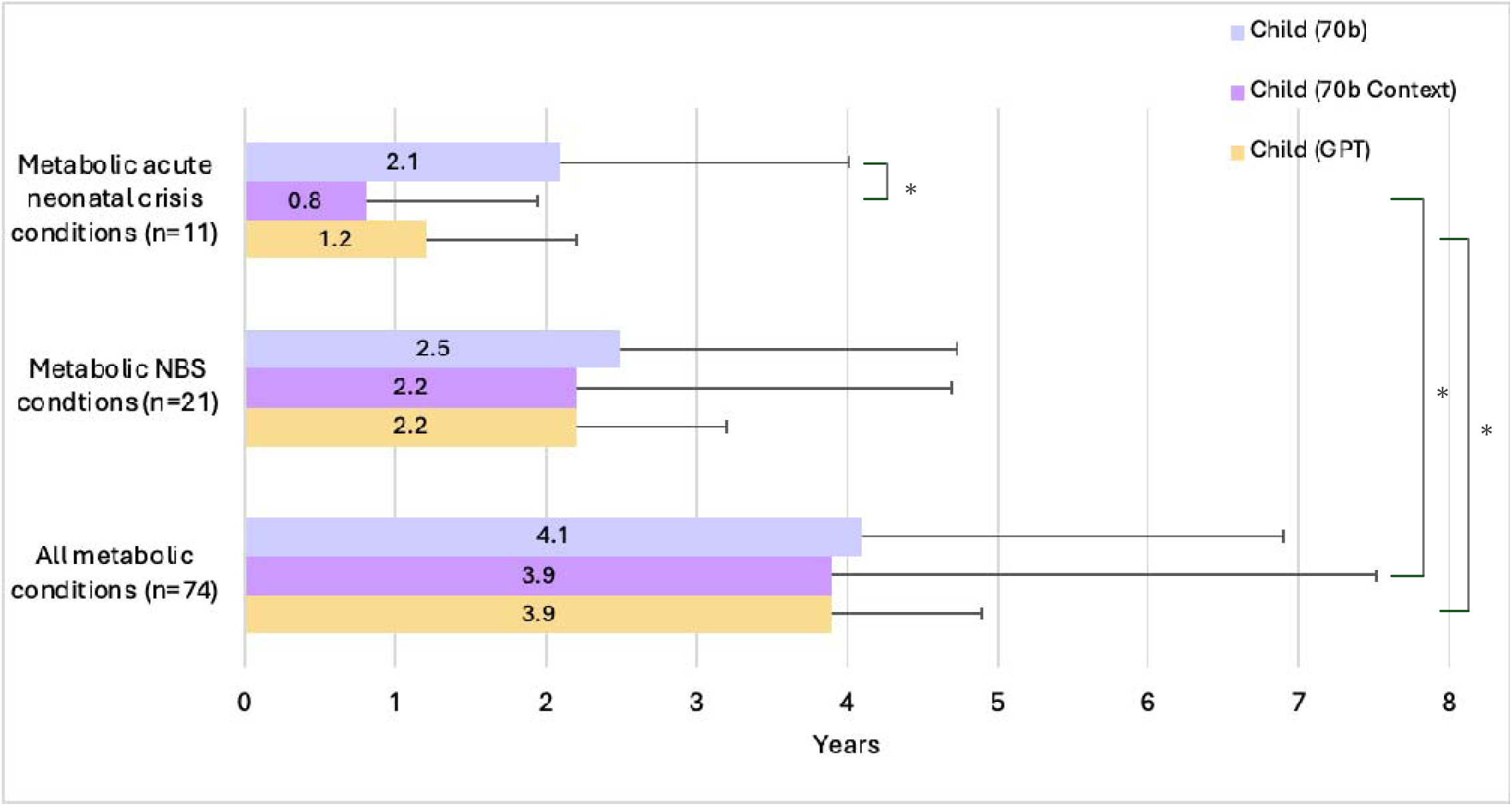
Mean child age presented in vignette output for metabolic conditions. Subgroupings of metabolic conditions were selected from the larger cohort of 282 conditions. The largest subgrouping contains “all metabolic conditions” within this study. The next grouping is only those “metabolic conditions listed within the RUSP for NBS”. The smallest grouping is that of “metabolic conditions with acute neonatal crisis”. Significance (*) is indicated at the α = 0.05 level, between Acute Neonatal Crisis conditions, Metabolic NBS conditions, and Metabolic conditions. The Bonferroni threshold for significance is p < 0.00566. NBS: Newborn Screening.

### Gender observations in LLM vignette output according to LLM and prompt type

From each vignette, gender signifiers such as “her” or “his” were used to assign gender to the described individual. Of the total 1692 generated vignettes, 27 did not have gender signifiers and instead used the wording, “the patient”. Compared to 70b and 70b Context, GPT is less likely to give gender signifiers for the child prompt with 23 of the total 282 (8.1%) vignettes lacking gender descriptions. Overall, gender ratios were heavily skewed toward male, particularly for 70b child prompts (7.3:1 male to female ratio; See Supplementary Table 7). Adult vignettes yielded a more even distribution of gender ratios (e.g, GPT child 4.4:1 compared to GPT adult 2.7:1). When context was applied in 70b, more female vignettes were generated, either lowering the male to female ratio for the child or skewing the adult male to female ratio more heavily female, 1.5:1 to 1:1.8. When removing those conditions that disproportionately affect one sex (Supplementary Table 8), we did not observe any significantly change in the male to female ratios. As expected, vignettes generated for conditions that disproportionately affect males demonstrated a male bias and those that disproportionately affect females overall showed a female bias, though not to the same degree as the male conditions (i.e., male to female ratios ranging from 3.9:1 to 32:1 for male conditions and 1:1 to 1:11 for female conditions (Supplementary Table 9). We emphasize that we extracted gender terms from the LLM output, and used those to assess conditions that may have different effects in individuals of different biological sexes, such as related to X-linked inheritance patterns.

### Mode of Inheritance of Conditions Included in Study

The Orphanet database served as the source for mode of inheritance for each of the 282 conditions in this study. Forty-five (16.0%) of the conditions had multiple modes of inheritance listed, whereas 229 (81.2%) had a single mode of inheritance and 8 (2.8%) are considered to typically occur sporadically. Supplementary Figure 3 shows the distribution of the modes of inheritance for the 282 conditions, with autosomal recessive being the most common inheritance pattern. To determine the impact of age of onset of a condition on the distribution of modes of inheritance, we compared “Limited to Childhood” conditions to “Limited to Adulthood” conditions (Supplementary Figure 4). Interestingly, there is a shift of the predominant mode of inheritance from autosomal recessive in “Limited to Childhood” to autosomal dominant in “Limited to Adulthood” conditions.

### Evaluating generated dialogues for the “Presentation Change” and “Management Change” categories

For the generated dialogues, we observed results with similar trends as the vignettes; that is, conditioned on any of the two studied categories “Presentation Change” or “Management Change”, the Correctness, Completeness, and Compassion scores were not statistically different with respect to age (p=0.352; p=0.548; p=0.903 for each of these performance metrics, respectively). The mean Total score was 13/15 (87%) and 12.9/15 (86%) for child dialogue and adult dialogue, respectively, which reveals a nonsignificant difference with respect to age (p=0.838) (see Table 1). Additionally, the open-source Llama-2-70b-chat demonstrated high performance in terms of quality of communication, with Compassion scores from 90-99% for all generated dialogues.

**Table 1.** Mean dialogue scores and statistical significance in Correctness, Completeness, Compassion, and Total metrics for dialogues generated in Management Change (n= 26) and Presentation Change (n= 15) with respect to age. Dialogues were only generated using Llama-2-70b-chat if the vignette for the associated condition and age group was accurate. Each dialogue was graded on a Likert scale, with 5 points available per metric and 15 total points available. Paired t-tests were conducted between the child and adult dialogues, conditioned on Correctness, Completeness, Compassion and Total scores. Significant differences at the α = 0.05 significance level are indicated with a marker (*), and all p-values were corrected using the Bonferroni correction (p < 0.00417).

|  | **Correctness**<br>**(0-5)** |  | **Completeness**<br>**(0-5)** |  | **Compassion**<br>**(0-5)** |  | **Total**<br>**(0-15)** |  |
| --- | --- | --- | --- | --- | --- | --- | --- | --- |
|  | **Child** | **Adult** | **Child** | **Adult** | **Child** | **Adult** | **Child** | **Adult** |
| Presentation<br>Change<br>(mean) | 3.93 | 4.07 | 4.27 | 4.27 | 4.93 | 4.93 | 13.1 | 13.3 |
| Presentation<br>Change<br>(p-value) | 0.743 |  | 1 |  | 1 |  | 0.857 |  |
| Management<br>Change<br>(mean) | 4.00 | 4.27 | 4.23 | 4.04 | 4.54 | 4.50 | 12.8 | 12.8 |
| Management<br>Change<br>(p-value) | 0.327 |  | 0.434 |  | 0.830 |  | 1 |  |
| All dialogues<br>(mean) | 4.20 | 3.98 | 4.12 | 4.24 | 4.67 | 4.68 | 13.0 | 12.9 |
| All dialogues<br>(p-value) | 0.352 |  | 0.548 |  | 0.903 |  | 0.838 |  |

### Evaluating generated management plans for the “Management Change” category

Unlike the observations for vignettes and dialogues, scores for management plans generated through Llama-2-70b-chat (n=53) showed lower results (See Table 2), ranging between 55-66% in terms of Correctness and Completeness, and ranging between 83-89% for Conciseness, with no statistical difference across different ages for Correctness, Completeness, and Conciseness (p= 0.642; p=0.182; p=0.371 for each of the three metrics respectively). This may be expected because we only provided the patient age and gender as in-context data and did not provide any other details about the diseases.

**Table 2.**
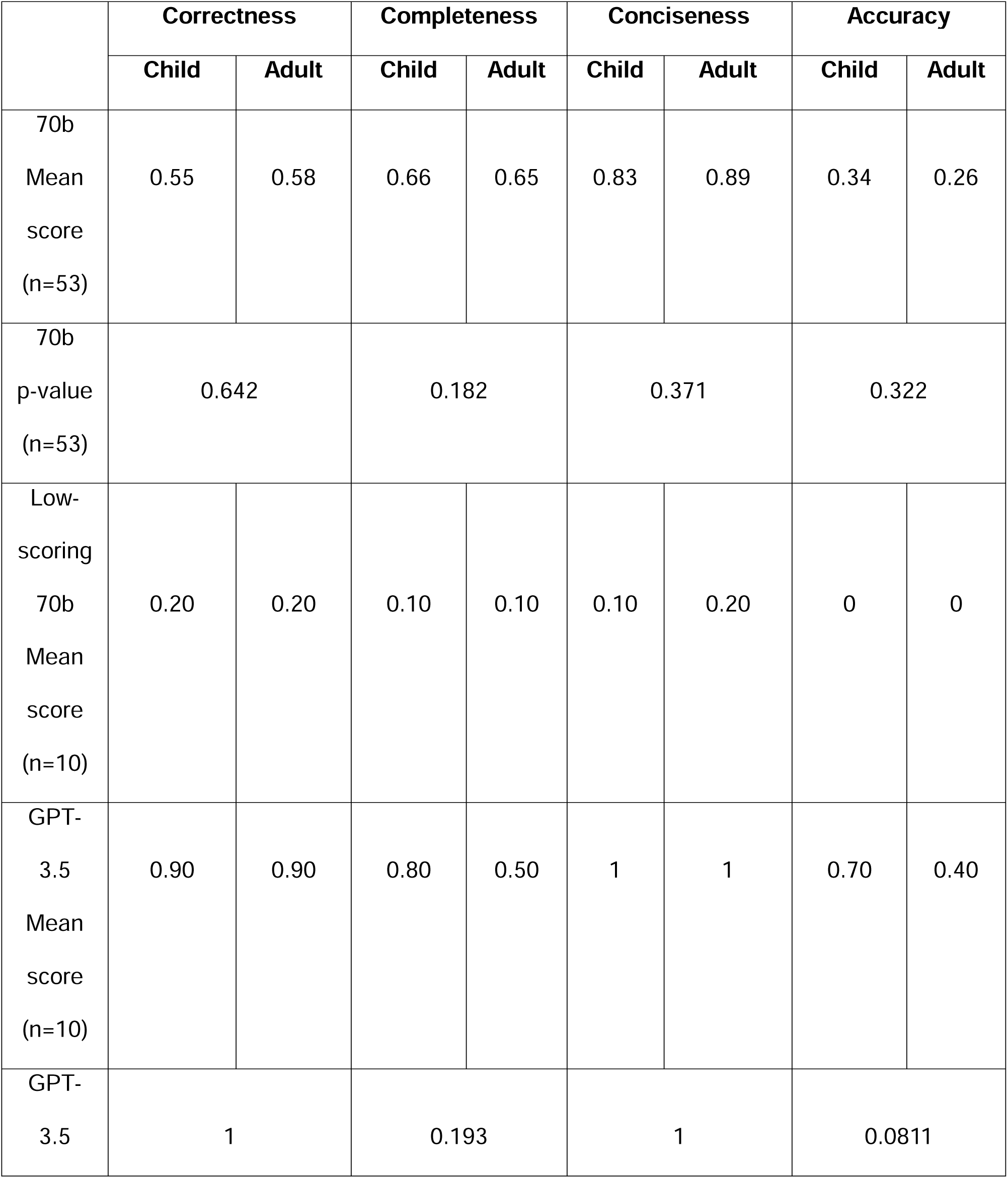
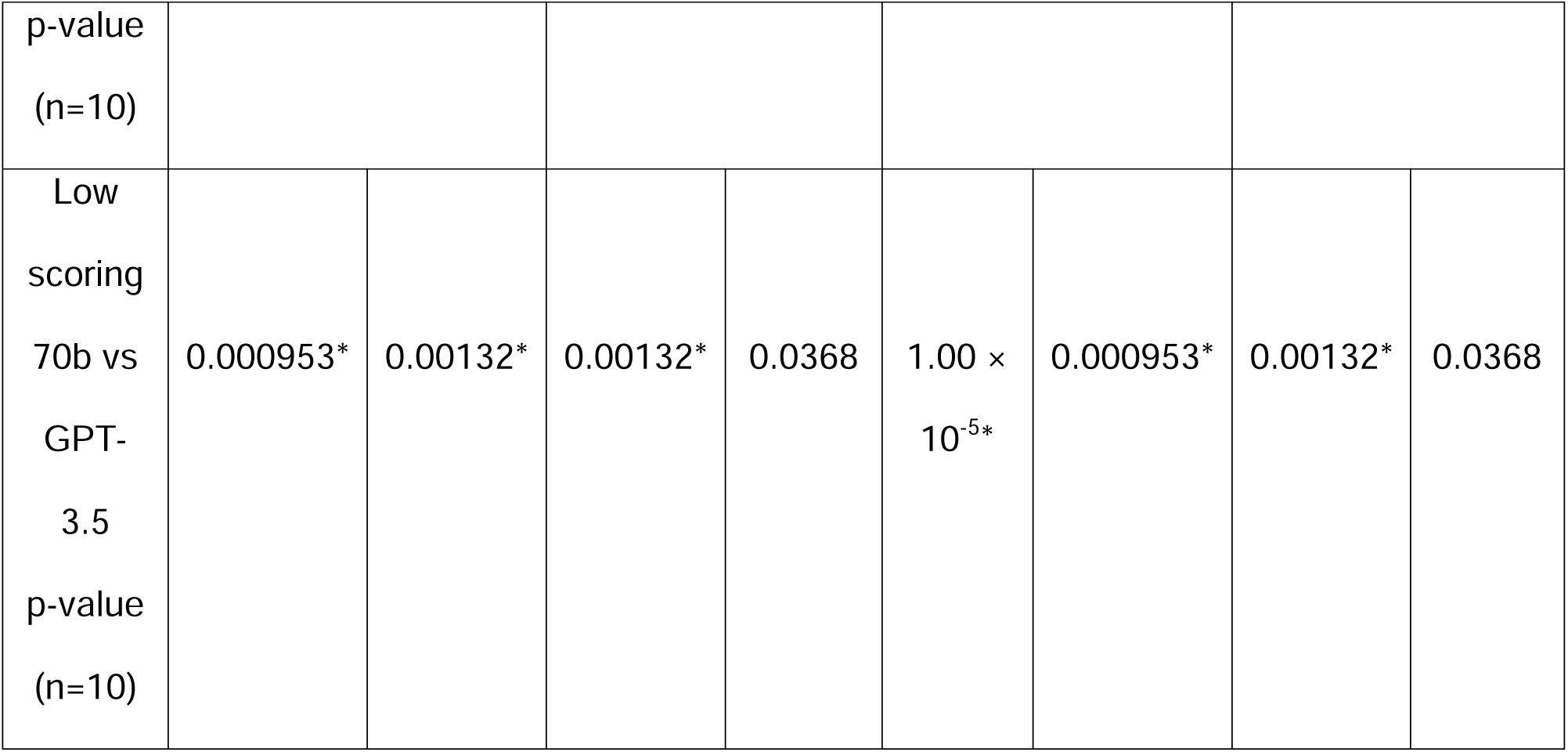
Summary of management scores and statistical comparison of child and adult management plans generated by Llama-2-70b-chat and GPT-3.5. Paired t-tests were conducted between the child and adult scores for Correctness, Completeness, Conciseness, and Accuracy of clinician-graded management plans generated by Llama-2-70b-chat (n = 53). Only management plans that scored a maximum of 1 out of 3 points using Llama-2-70b-chat were repeated using GPT-3.5 (n = 10). Because of very low sample size (n=10), permutation tests were conducted between the child and adult scores in Correctness, Completeness, Conciseness, and Accuracy of clinician-graded management plans generated by GPT-3.5. Permutation tests were also conducted between the low-scoring Llama-2-70b-chat entries and GPT-3.5 for Correctness, Completeness, Conciseness and Accuracy scores in child and adult age groups (n = 10). Significant differences at the α = 0.05 significance level are indicated with a marker (*), and all p-values were corrected using the Bonferroni correction (p-value threshold = 0.00833).

Since the three individual performance metrics were low, the Accuracy score was also low, ranging from 26%-34%, with no statistically significant difference between child and adult age groups (p=0.322). GPT was used to generate child and adult management plans for the conditions that scored low on 70b, which showed statistically significant improvement in Correctness for child plans (p=0.000953) and adult plans (p=0.00132). Completeness scores for GPT-generated child and adult plans improved compared to the low 70b scores (child: 10% to 80%; adult: 10% to 50%). The improvement in child plans is significant (p=0.00132) while the improvement in adult plans is not statistically significant. This non-significant difference for the adult plans could be attributed to significantly higher Conciseness score for GPT that might have resulted in lower Completeness scores as a consequence of presenting the information concisely. As a result of this non-significant improvement for GPT for the adult plans, the adult Accuracy score did not show an overall significant improvement (p=0.0368). Conversely, for the child group, GPT resulted in significantly improved Accuracy (p=0.00132).

## Discussion

Language models can produce unintentional age or gender-related biases in their responses simply because of disproportionally distributed training datasets with respect to the source for these biases.^26,27^ This study seeks to examine whether age-biases exist in LLM responses with respect to genetic conditions whose manifestations and management plans may change with respect to age. In our study, the tested LLM models were Llama-2-70b-chat and GPT-3.5, and the kinds of generated outputs were the medical vignettes, patient-doctor dialogues, and management plans. The evaluation was done with respect to 282 prevalent genetic conditions and a subgroup of 74 metabolic conditions. Very rare disorders were excluded from this study, and we plan to expand the number of diseases in future work.

Our original assumption was that, for most conditions, due to the focus on pediatric presentations of disease, including related to importance of early diagnosis and intervention, we would observe an effect of the preponderance of written records describing clinical findings and treatment plans for the child group versus the adult group. Hence, we originally expected LLM responses to be more realistically plausible at generating medical vignettes, patient-doctor dialogues, and management plans for the child group than for the adult group. To our surprise, Llama-2-70b-chat (with and without in-context prompting) and GPT-3.5 (without in-context prompting) did not show any obvious age-related biases.

For the medical vignettes, among the diseases in the categories “Presentation Change”, “Management Change”, and “No Change”, Figures 5-8 and Supplementary Figures 1 and 2 did not show statistically differences in the Correctness and Completeness scores between the child and adult groups. LLM only fails at generating medical vignettes of an adult patient for diseases in “Limited to Childhood” (and likewise, child vignettes for diseases in “Limited to Adulthood”). However, this behavior is fully expected.

Besides being unaffected with obvious age-biases, 70b (with in-context prompting) and GPT (without in-context prompting) both obtain a surprisingly high performance at generating medical vignettes; both Correctness and Completeness score were at least 0.68 out of 1, with most scores being close to 0.8 out of 1, averaging over 282 diseases (Supplementary Figures 1 and 2).

These findings suggest that, contrary to our initial suspicions, 70b Context and GPT were well-trained with respect to age distribution and thus produce plausible descriptions of disease manifestations (e.g., vignettes) for both the child and adult group. This is particularly helpful due to the lack of formal genetics training/curriculum in many internal medicine and primary care residency programs despite the increasing need and interest among those trainees in genetics as they continue to take care of more adult individuals with an underlying genetic condition, and as more people with genetic conditions survive into adulthood or are diagnosed later in life.^28–31^ Nowadays, many of these clinical trainees, as well as others, use those LLMs on a frequent basis whether during their clinical service or while studying for medical exams and board certifications to retrieve information and learn more about the clinical features, mode of inheritance, and outcomes of these genetic conditions.^32–34^

We noted that the mean age of a child averaging over all the generated child vignettes was 5 ±3.4 years. We also evaluated LLM performance on the child vignettes conditioned only on metabolic diseases (Figure 10). Certain kinds of metabolic diseases are expected to manifest at different stages of life. We considered three groups of metabolic diseases (1) “All metabolic conditions” contained within our original cohort; (2) Sub-group of “metabolic conditions listed within RUSP for NBS” and (3) A smaller sub-group of “metabolic conditions with acute neonatal crisis” where these acute events are a very important medical issue (e.g., MSUD). Results showed a statistically significant decrease in mean age generated by 70b Context of 3.9 years versus 0.8 years between “all metabolic conditions” group versus the group of “metabolic conditions with acute neonatal crisis” (p= 0.00648). With GPT, more statistically significant results were seen, with mean age being 3.9 years vs. 1.2 years between these two disease sets, respectively (p= 0.00226). This likely reflects differences in training data and may points towards the knowledge of these LLM models about the very early onset of these “metabolic conditions with acute neonatal crisis” which tend to present soon after birth. This significant difference in mean age was not observed when comparing “all metabolic conditions” group to the group of “metabolic conditions listed within RUSP for NBS”, possibly because several of these conditions have insidious onset and if treated early in life, can result in an increased frequency of later-onset information related to longer-term sequelae (e.g., for Phenylketonuria and Homocystinuria). Thus, when we examine subsets of disease, we can see that the LLMs assign ages that correlate with clinical expectations based on important medical issues.

We also analyzed the mode of inheritance in the generated medical vignettes with respect to the 282 selected conditions. This analysis does not strictly address age-related biases; however, we found it to be an interesting question to help us understand how genetic conditions were described related to another important area in medical genomics. We found that there was a shift in inheritance mode, going from autosomal recessive in “Limited to Childhood” to autosomal dominant in “Limited to Adulthood” conditions (Supplementary Figure 4). A possible explanation for this could be that autosomal recessive conditions tend to be more severe compared to autosomal dominant conditions. This is likely due to evidence that disease genes that involve recessive inheritance are under different selective pressures than those that involve dominant inheritance and thus may, in general, manifest in diseases at different ages (and become “Limited to Childhood”). One example is the Autosomal Recessive Polycystic Kidney Disease (ARPKD) which typically presents much earlier and is more severe than Autosomal dominant polycystic kidney disease (ADPKD).^35–37^ We note that this is a generalization and does not take into account scenarios such as related to deleterious de novo variants.

From the generated vignettes, we further generated dialogues between a hypothetical clinical geneticist and a hypothetical patient using these vignettes as in-context prompts. This simulation aims to reveal some insights about how an actual patient may converse with the LLM. Overall, Llama-2-70b-chat could generate realistically plausible dialogues with a mean Total score of 13/15 (87%) and 12.9/15 (86%) for the child and adult group, respectively (Table 1). In terms of quality of communication, the Compassion scores excel the most, ranging from 4.5-4.93/5 (90-99%) for all generated dialogues. This is especially important given the potential use of these LLMs and chatbots for conversations by patients as well as physicians, including to draft responses to patients or medical colleagues; however further exploration of this technology is warranted given the high-risk nature of clinical communication. In this paper, we only graded each generated dialogue in its entirety; that is, we focused on how plausible the entire dialogue appears. However, some issues emerge upon close observation. For example, there can be a robotic nature to these generated dialogues, which would hopefully not occur in actual patient-doctor conversations. Future study will focus on analysis for these types of nuances.

Unlike the generated vignettes and dialogues, management plans generated by Llama-2-70b-chat obtained low Correctness and Completeness scores (55-66%), but better Conciseness (83-89%) for the child and adult age group (Table 2). One potential explanation might be that the Llama-2-70b-chat was not provided with comprehensive in-context information, but rather only given the patient’s age and gender as in-context prompts. When GPT-3.5 was used to generate management plans, Accuracy score significantly improved for the child group (p=0.00132) but showed a smaller, non-significant improvement for the adult group (p=0.0368) with multiple testing correction. These results in general support that these LLM models can encode a wealth of semantic knowledge about genetic conditions and have high conversational abilities with quality communications but are still not ready or safe for critical decisions related to management plans, including as they lack real-world implementation evidence.^38,39^ We emphasize the following main difference between the management plans and the dialogues.

Although dialogues were also assessed for the appropriateness of their management recommendations conveyed to the patient during the conversation, the management recommendations were assessed from a more general viewpoint and with different expectations. The dialogues described an initial evaluation during a first encounter between the patient and the geneticist and focused on discussing possible expected diagnoses, possible clinical outcomes, and initial recommendations including sending testing to confirm suspected diagnoses, referring to other subspecialists, and scheduling follow-up visits to review testing results and provide more specific recommendations in subsequent visits. Conversely, when generating management plans, the LLMs were asked specific management questions for a specific known diagnosis, and therefore a more comprehensive and specific output was expected.

We note that the vignettes were manually analyzed more extensively than the dialogues and management plans. Our rationale was that the vignettes were used as in-context prompting for the dialogues. Age and gender of the hypothetical patient in the vignettes were used for generating management plans. Our future study would focus on more detailed analysis of the dialogues and management plans. For example, suppose certain phenotypes were excluded during the conversation, then we would evaluate how the final disease diagnosis may change. For management plans, we could include or exclude certain test results and then observe how the LLM response may appear. Moreover, we generated vignettes and dialogues with respect to two broad age groups, child and adult. In future work, we would further focus on narrower age categories, where vignettes, dialogues, and management plans are generated and graded with respect to many age brackets (e.g., neonatal, infancy, childhood, adolescence, adult, elderly).

On the topic of model limitation, we note that we used Llama-2-70b-chat and GPT-3.5 without editing any default parameters like top_k and top_p, which can affect the response generation process. Moreover, we note that one could finetune LLM specifically on medical datasets or on the generated vignettes and dialogues.^15^ This would require a large amount of well curated data (either automatically or manually). For example, to finetune their model, Tu et al generated and auto-evaluated many hypothetical vignettes and dialogues, though most of which are not geared specifically towards genetic conditions with potential age-related manifestations and management plans. However, as mentioned previously, we encountered both computational cost and annotation problems when following the Tu et al auto-evaluation protocol (e.g., few-shot in-context learning).

However, in many situations, the average users would not finetune the LLM with respect to their own datasets. Rather, we suspect that the average users would rely on useful prompts to guide LLM output. For these reasons, in this paper, we evaluated the performance of only pre-trained LLMs on diseases whose manifestations and management may differ with respect to age. Our approach is likely to reveal more plausible insights about how LLMs are being used by both clinicians and patients regarding these genetic conditions in practice.

On the topic of in-context prompting, this strategy continues to be effective in improving the performance of LLMs in a specific knowledge area.^23^ The primary limitation with in-context prompting is the token limit size of each LLM. Llama-2-70b-chat has a token size limit of 4096 tokens; after incorporating information from both Orphanet and GeneReviews, the token limit is almost reached. Other studies have investigated the use of bulk scientific literature (including online searches outside of medical databases) to guide their own LLMs.^15^ However, due to computational limitations and token limits of open-source Llama-2-70b-chat, we were unable to incorporate much more than Orphanet and GeneReviews descriptions. Despite this, Llama-2-70b-chat still performed impressively compared to GPT-3.5 at generating medical vignettes considering the model size difference (see Supplementary Tables 5 and 6). In future work, we plan to deploy the experiment based on real-life clinical notes and/or conversations, where there can be much more variability among the input context for the same disease. Such settings would enable the LLM generated vignettes and dialogues to be much more diverse.

The token limit of a LLM model can also affect the complexity of the data generation process. For example, previous work implemented a “critic agent” that knows the ground-truth and provides feedback to the “doctor agent” after the first chat session.^15^ This feedback and all the first chat session data would then be used as an in-context prompt for the second chat session. Hence, the second chat session would tend to provide higher quality dialogues. With the token limit of Llama-2-70b-chat, we were only able to implement the “patient” and “doctor” agent without the “critic” (i.e., without feedback to improve the “doctor agent” in the second chat session). Moreover, we could only generate the dialogues for one single chat session to avoid using all the tokens.

In summary, the tested LLMs exceeded expectations in addressing genetic conditions, even in adult scenarios. However, it is important to highlight that despite these impressive capabilities, at least open-source LLMs (and likely all LLMs) still have significant limitations in providing age-related management decisions, which should be taken with significant caution given the wide use of these LLMs by patients, medical providers and providers in training.

## Author contributions statement

AAO, KAF, RLW, DD, and BDS were responsible for conceiving and conducting experiments, analyzing the results, and writing and reviewing the manuscript. SLH and PH were responsible for conducting experiments, analyzing the results, and reviewing the manuscript.

## Acknowledgements

This research was supported by the Intramural Research Program of the National Human Genome Research Institute, National Institutes of Health. This work utilized the computational resources of the NIH HPC Biowulf cluster.

