## Supplementary Figures and Tables for "Assessing Large Language Model Performance Related to Aging in Genetic Conditions"

**Supplementary Table 1. For the following 6 diseases, Roberta embedding was used to compute the cosine similarity score between each of the 10 repeated vignette generations with the original vignette.** Similarity score ranges from -1 to 1, with 1 as the most similar. In the last row, one additional comparison was made (Tibial Muscular Dystrophy- child vs Fryns syndrome- adult) to demonstrate what the similarity scores are for two dissimilar vignettes. To further provide context about these similarity scores, we also computed the score between two highly similar sentences: “I like ice cream” and “I love ice cream” and found the similarity score to be 0.891.

|  | 1 | 2 | 3 | 4 | 5 | 6 | 7 | 8 | 9 | 10 | Average |
| --- | --- | --- | --- | --- | --- | --- | --- | --- | --- | --- | --- |
| Tibial Muscular Dystrophy (child) | 0.712 | 0.865 | 0.633 | 0.914 | 0.636 | 0.564 | 0.691 | 0.948 | 0.711 | 0.780 | 0.745 |
| Tibial Muscular Dystrophy (adult) | 0.912 | 0.943 | 0.921 | 0.898 | 0.909 | 0.941 | 0.934 | 0.919 | 0.950 | 0.927 | 0.925 |
| Amyotrophic lateral sclerosis (child) | 0.789 | 0.843 | 0.811 | 0.803 | 0.774 | 0.791 | 0.613 | 0.789 | 0.807 | 0.648 | 0.767 |
| Amyotrophic lateral sclerosis (adult) | 0.860 | 0.908 | 0.799 | 0.650 | 0.884 | 0.909 | 0.855 | 0.871 | 0.881 | 0.841 | 0.846 |
| Leber hereditary optic neuropathy (child) | 0.804 | 0.744 | 0.744 | 0.781 | 0.753 | 0.640 | 0.754 | 0.750 | 0.718 | 0.796 | 0.748 |
| Leber hereditary optic neuropathy (adult) | 1.000 | 1.000 | 0.831 | 0.898 | 0.920 | 0.894 | 0.905 | 0.932 | 0.929 | 0.862 | 0.917 |
| Aicardi-Goutières syndrome (child) | 0.794 | 0.794 | 0.850 | 0.901 | 0.932 | 0.935 | 0.898 | 0.898 | 0.915 | 0.930 | 0.885 |
| Aicardi-Goutières syndrome (adult) | 0.951 | 0.836 | 0.906 | 0.808 | 0.832 | 0.922 | 0.914 | 0.928 | 0.918 | 0.940 | 0.896 |
| Fryns syndrome (child) | 0.802 | 0.711 | 0.887 | 0.830 | 0.672 | 0.797 | 0.832 | 0.718 | 0.752 | 0.851 | 0.785 |
| Fryns syndrome (adult) | 0.912 | 0.844 | 0.833 | 0.882 | 0.868 | 0.900 | 0.891 | 0.851 | 0.918 | 0.828 | 0.873 |
| Meckel syndrome (child) | 0.782 | 0.812 | 0.683 | 0.782 | 0.717 | 0.814 | 0.672 | 0.746 | 0.835 | 0.821 | 0.767 |
| Meckel syndrome (adult) | 0.706 | 0.804 | 0.733 | 0.746 | 0.715 | 0.721 | 0.756 | 0.747 | 0.808 | 0.752 | 0.749 |
| Tibial Muscular Dystrophy (child) vs Fryns (adult) | 0.407 | 0.415 | 0.484 | 0.446 | 0.401 | 0.417 | 0.609 | 0.413 | 0.440 | 0.509 | 0.454 |

**Supplementary Table 2: Scoring rubric for the medical vignettes and management plans.** We assessed all vignette (n=282) and management responses (n=53) for Correctness, Completeness, and Conciseness, assigning a score of either 0 or 1 for each metric. For the medical vignettes, since all results received Conciseness score of 1, we removed this metric when evaluating the total score of the vignettes, thus allowing a maximum total score of 2 for each vignette. For management results, Conciseness was still included in the analysis, yielding a maximum Total score of 3 for each management answer.

| **Criteria** | **Score** | **Definition** |
| --- | --- | --- |
| **Correctness*** | 0 | Inaccurate and inappropriate information or description of manifestations not relevant to the genetic disorder, the patient’s age, or gender. |
|  | 1 | Accurate and appropriate description of manifestations (presented without errors) of the genetic disorder considering the patient’s age and gender. |
| **Completeness*** | 0 | Missing crucial patient information or cardinal clinical or laboratory features of the genetic disorder. |
|  | 1 | Provides a complete list of important clinical or laboratory features relevant to the genetic disorder.** |
| **Conciseness** | 0 | Includes unnecessary or repetitive information or lacks focus on key points. |
|  | 1 | Clearly and concisely presents relevant information without unnecessary or repetitive details. |

***** A separate metric “Accuracy” was marked as 1 only if a vignette received a score of 1 for both correctness and completeness.

** Considering that not all individuals affected by a genetic condition display all the textbook characteristics, but instead demonstrate varying degrees of severity, clinicians evaluating the vignettes considered a vignette complete if it exhibited the most essential features along with sufficient additional clinical and laboratory indicators for a given condition.

**Supplementary Table 3. Kappa statistic to evaluate the agreement among the clinicians while grading the vignettes for Correctness and Completeness.** Kappa statistic close to 1 indicates the highest agreement, whereas 0 indicates no agreement.

|  | Clinician 1 & Clinician 2 | Clinician 1 & Clinician 3 | Clinician 2 & Clinician 3 |
| --- | --- | --- | --- |
| Kappa statistic (Correctness) | 0.6 | 0.61 | 0.74 |
| Kappa statistic (Completeness) | 0.52 | 0.55 | 0.88 |

**Supplementary Table 4: Scoring rubric for patient-geneticist dialogues on 5-point Likert scale.** Since a dialogue is longer than a vignette and management plan, each of the three metrics was graded on a more comprehensive 5-point Likert scale to more precisely assess differences. Compassion metric was added to the patient-geneticist dialogue rubric to further evaluate the quality of communication provided by the LLMs.

| **Criteria** | **Definition** | **1 (Strongly Disagree)** | **2 (Disagree)** | **3 (Uncertain)** | **4 (Agree)** | **5 (Strongly Agree)** |
| --- | --- | --- | --- | --- | --- | --- |
| **Correctness** | The dialogue contains **realistic**, accurate and appropriate descriptions (or management) relevant to the genetic disorder, the patient’s age, or gender. | Contains numerous errors, significantly impacting understanding. | Contains several errors, affecting understanding. | Contains occasional errors, slightly affecting understanding | Contains few errors but generally does not affect understanding of key concepts. | Contains no errors, greatly enhanced understanding. |
| **Completeness** | The dialogue provides a complete list of important clinical, laboratory features [HPO terms], and management options relevant to the genetic disorder | The dialogue lacks crucial information, leaving significant gaps in understanding and treatment planning. | The dialogue is missing some important details, leading to gaps in understanding and treatment planning. | The dialogue includes most relevant information but may lack some minor details. | The dialogue includes nearly all relevant information, facilitating comprehensive understanding and treatment planning. | The dialogue is very thorough, providing all necessary information for comprehensive understanding and treatment planning. |
| **Compassion** | The dialogue shows empathy, addresses concerns, provides reassurance, asks open-ended questions, provides resources, avoids jargon, and checks understanding. | Strongly Disagree | Disagree | Neither agree nor disagree | Agree | Strongly Agree |

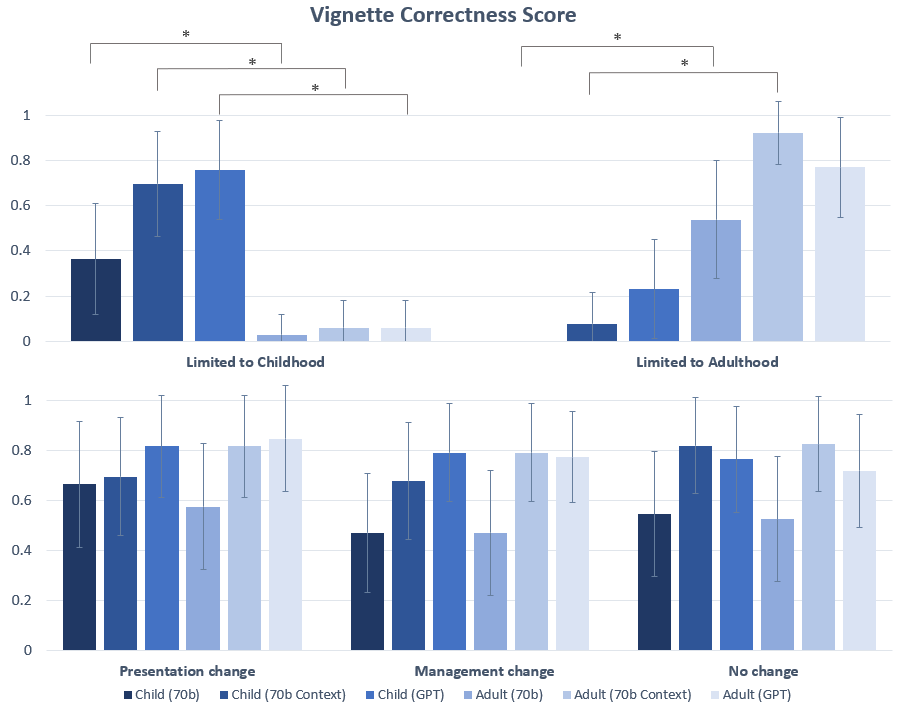

**Supplementary Figure 1. Individual Correctness score breakdown for medical vignettes with error and significance in all 5 categories (Limited to Childhood, Limited to Adulthood, Presentation Change, Management Change, and No Change).** All vignettes were clinician-graded and assigned a score of either 0 or 1 for Correctness. One asterisk (*) indicates significance between child and adult vignettes for the Correctness score. In this chart, significance is only demonstrated for comparisons between child and adult vignettes for the same model (see Supplementary X for statistical comparisons between 70b, 70b Context and GPT).

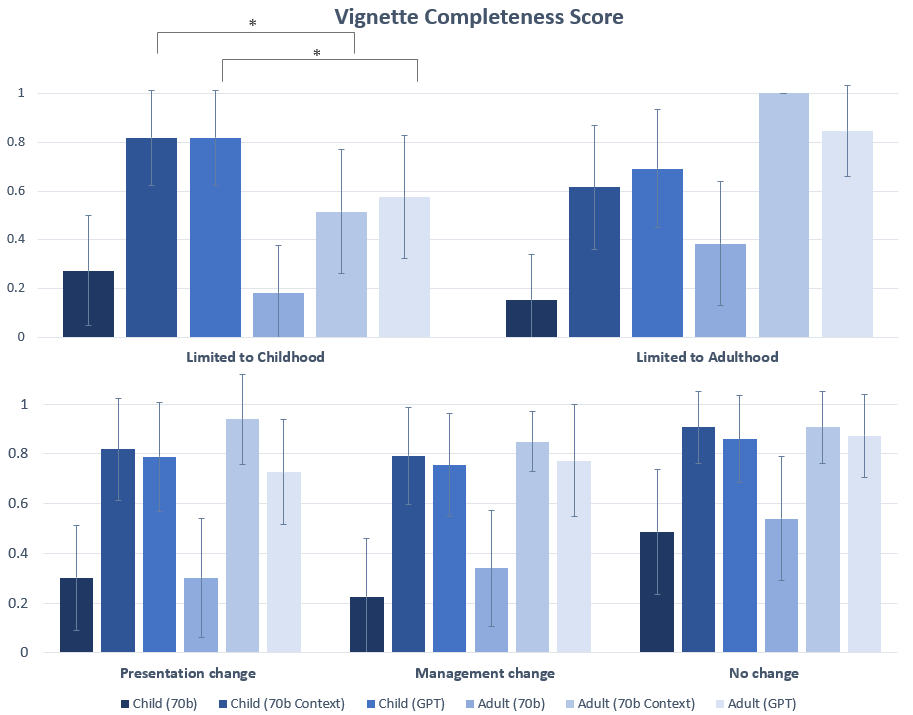

**Supplementary Figure 2. Individual Completeness score breakdown for medical vignettes with error and significance for all 5 categories (Limited to Childhood, Limited to Adulthood, Presentation Change, Management Change, and No Change).** All vignettes were clinician-graded and assigned a score of either 0 or 1 for Completeness. One asterisk (*) indicates significance between child and adult vignettes for the Completeness score. In this chart, significance is only demonstrated for comparisons between child and adult vignettes for the same model (see Supplementary X for statistical comparisons between 70b, 70b Context and GPT).

**Supplementary Table 5. Statistical comparison of llama-2-70b-chat vignette scores with and without in-context prompting (70b vs 70b Context)**. Paired t-tests were conducted between the Correctness, Completeness, and Accuracy scores of clinician-graded medical vignettes.

| **Category** | **Correctness (p-value)** | | **Completeness (p-value)** | | **Accuracy (p-value)** | |
| --- | --- | --- | --- | --- | --- | --- |
|  | **Child** | **Adult** | **Child** | **Adult** | **Child** | **Adult** |
| Limited to childhood | 0.00296 | 0.325 | 1.58 × 10^-5^* | 0.00296 | 0.00300 | 0.325 |
| Limited to adulthood | 0.337 | 0.0180 | 0.00753 | 0.000893* | 0.337 | 0.00281 |
| Presentation change | 0.744 | 0.0184 | 1.02 × 10^-5^* | 1.61 × 10^-8^* | 0.00123* | 1.77 × 10^-6^* |
| Management change | 0.0329 | 0.000371* | 4.28 × 10^-10^* | 3.86 × 10^-8^* | 0.252 | 0.0586 |
| No Change | 2.23 × 10^-8^* | 1.40 × 10^-11^* | 8.66 × 10^-17^* | 7.76 × 10^-13^* | 3.41 × 10^-15^* | 6.12 × 10^-17^* |

*The Bonferroni threshold for significance is p<0.00167.

**Supplementary Table 6. Statistical comparison of llama-2-70b-chat vignette scores with in-context prompting to GPT-3.5 vignette scores (70b Context vs GPT).** Paired t-tests were conducted between the Correctness, Completeness, and Accuracy scores of clinician-graded medical vignettes. None of these values are significant at the α = 0.05 significance level using the Bonferroni correction (p < 0.00167).

| **Category** | **Correctness (p-value)** | | **Completeness (p-value)** | | **Accuracy (p-value)** | |
| --- | --- | --- | --- | --- | --- | --- |
|  | **Child** | **Adult** | **Child** | **Adult** | **Child** | **Adult** |
| Limited to Childhood | 0.587 | 1 | 1 | 0.627 | 0.436 | 1 |
| Limited to Adulthood | 0.298 | 0.298 | 0.695 | 0.165 | 0.298 | 0.150 |
| Presentation Change | 0.257 | 0.746 | 0.761 | 0.0215 | 0.608 | 0.179 |
| Management Change | 0.190 | 0.816 | 0.646 | 0.325 | 0.319 | 0.151 |
| No Change | 0.255 | 0.0274 | 0.209 | 0.358 | 0.126 | 0.0117 |

**Supplementary Table 7. Gender bias in LLM vignette output according to LLM and prompt type**. Of the 282 conditions in this study, 34 are known to preferentially affect males and 12 are known to preferentially affect females (based on source data from Orphanet: https://www.orpha.net/en/disease). However, removing conditions with a known gender predilection did not ameliorate the gender bias produced by the LLMs. Gender output was more equitable in the adult prompted vignettes compared to the child for both LLM types. Interestingly, when context was provided to the 70b prompt, gender ratio screwed more heavily female for adult vignettes. Of note, GPT did not provide gender for 23 (8.1%) of the child vignettes, whereas in the adult prompt it did not provide gender for just 2 vignettes. Gender was given in 70b generated vignettes except for 1 in Adult (70b) and Child (70b Context).

|  | **All Conditions**  **(n=282)** | | | **Conditions that affect**  **males and females proportionately**  **(n=236)** | | |
| --- | --- | --- | --- | --- | --- | --- |
|  | Male (M) | Female (F) | M:F Ratio | Male (M) | Female (F) | M:F Ratio |
| **Child (70b)** | 248 | 34 | 7.3:1 | 210 | 26 | 8.1:1 |
| **Child (70b Context)** | 234 | 47 | 5:1* | 199 | 36 | 5.5:1* |
| **Child (GPT)** | 211 | 48 | 4.4:1* | 174 | 41 | 4.2:1* |
| **Adult (70b)** | 167 | 114 | 1.5:1* | 134 | 101 | 1.3:1* |
| **Adult (70b Context)** | 100 | 82 | 1:1.8 | 72 | 164 | 1:2.3 |
| **Adult (GPT)** | 205 | 75 | 2.7:1* | 171 | 64 | 2.7:1 |

*Gender was not given for all vignettes.

**Supplementary Table 8. Conditions in this study that disproportionately affect males or females.** The Orphanet database (<https://www.orpha.net/en/disease>) served as the source of information about sex specific prevalence unless otherwise noted by the superscript letters and citations below.

| **Conditions that disproportionately affect Males (n=34)** | | **Conditions that disproportionately affect Females (n=12)** |
| --- | --- | --- |
| Aarskog-Scott syndrome | Mucopolysaccharidosis type 2 | Acute intermittent porphyria |
| Barth syndrome | Oculocerebrorenal syndrome of Lowe | Crouzon syndrome-acanthosis nigricans syndrome |
| Brugada syndrome | Ornithine transcarbamylase deficiency | Currarino syndrome |
| Chronic granulomatous disease | Paroxysmal non-kinesigenic dyskinesia | Incontinentia pigmenti |
| Complete androgen insensitivity syndrome | Pelizaeus-Merzbacher disease | Orofaciodigital syndrome type 1 |
| Duchenne muscular dystrophy | Recessive X-linked ichthyosis | Porphyria variegate |
| Fabry disease | Retinitis pigmentosa | Pseudoxanthoma elasticum |
| Fragile X syndrome | Shwachman-Diamond syndrome^b^ | Rett syndrome |
| Glycogen storage disease due to liver phosphorylase kinase deficiency^a^ | Syndromic recessive X-linked ichthyosis | Trisomy 18 |
| Hemophilia A | Triploidy^c^ | Turner syndrome |
| Hemophilia B | Wiskott-Aldrich syndrome | Wolf-Hirschhorn syndrome |
| Hypohidrotic ectodermal dysplasia | X-linked adrenal hypoplasia congenita | X-linked dominant chondrodysplasia punctata |
| Kallmann syndrome | X-linked agammaglobulinemia |  |
| Leber hereditary optic neuropathy | X-linked centronuclear myopathy |  |
| Leigh syndrome | X-linked Emery-Dreifuss muscular dystrophy |  |
| Lesch-Nyhan syndrome | X-linked recessive ocular albinism |  |
| Menkes disease | X-linked retinoschisis |  |

a. Kishnani PS, Goldstein J, Austin SL, Arn P, Bachrach B, Bali DS, Chung WK, El-Gharbawy A, Brown LM, Kahler S, Pendyal S, Ross KM, Tsilianidis L, Weinstein DA, Watson MS; ACMG Work Group on Diagnosis and Management of Glycogen Storage Diseases Type VI and IX. Diagnosis and management of glycogen storage diseases type VI and IX: a clinical practice resource of the American College of Medical Genetics and Genomics (ACMG). Genet Med. 2019 Apr;21(4):772-789. Doi: 10.1038/s41436-018-0364-2. Epub 2019 Jan 19. PMID: 30659246.
b. Farooqui SM, Ward R, Aziz M. Shwachman-Diamond Syndrome. [Updated 2023 Jul 17]. In: StatPearls [Internet]. Treasure Island (FL): StatPearls Publishing; 2024 Jan-. Available from:
c. Kolarski M, Ahmetovic B, Beres M, Topic R, Nikic V, Kavecan I, Sabic S. Genetic Counseling and Prenatal Diagnosis of Triploidy During the Second Trimester of Pregnancy. Med Arch. 2017 Apr;71(2):144-147. Doi: 10.5455/medarh.2017.71.144-147. PMID: 28790549; PMCID: PMC5511524.

**Supplementary Table 9. Vignette gender output for conditions that disproportionately affect males or females**. Of the 282 conditions in this study, 34 are known to preferentially affect males and 12 are known to preferentially affect females. The majority of the male conditions (22/34) can affect females, some rarely (e.g., Wiskott-Aldrich syndrome) and some with a more minor disproportionate ratio (e.g., Paryoxysmal non-kinesigenic dyskinesia, 1.4 males affected for each female). For females, only 2 out of the 12 conditions are known to exclusively affect females (Rett and Turner syndrome), whereas the other conditions vary in sex ratios.

|  | **Conditions that disproportionately affect Males**  **(n=34)** | | | **Conditions that disproportionately affect Females**  **(n=12)** | | |
| --- | --- | --- | --- | --- | --- | --- |
|  | Male (M) | Female (F) | M:F Ratio | Male (M) | Female (F) | M:F Ratio |
| **Child (70b)** | 32 | 2 | 16:1 | 6 | 6 | 1:1 |
| **Child (70b Context)** | 31 | 3 | 10.3:1 | 4 | 8 | 1:2 |
| **Child (Chat GPT)** | 33 | 1 | 33:1 | 4 | 6 | 1:1.5* |
| **Adult (70b)** | 32 | 2 | 16:1 | 1 | 11 | 1:11 |
| **Adult (70b Context)** | 27 | 7 | 3.9:1 | 1 | 11 | 1:11 |
| **Adult (Chat GPT)** | 32 | 1 | 32:1 | 2 | 10 | 1:5 |

*Gender was not provided for 2 vignettes.

|  | Number of conditions |
| --- | --- |
| Autosomal recessive | 169 |
| Autosomal dominant | 112 |
| X-linked recessive | 35 |
| X-linked dominant | 9 |
| Mitochondrial | 5 |
| Sporadic | 8 |

**Supplementary Figure 3. Mode of inheritance for the 282 conditions included in this study.** Forty-five of these conditions have multiple modes of inheritance. Therefore, a single condition could represent multiple points. In addition to hereditary conditions, there are 8 conditions that are typically sporadic.

|  | Autosomal recessive | Autosomal dominant | X-linked recessive | X-linked dominant | Mitochondrial | Sporadic |
| --- | --- | --- | --- | --- | --- | --- |
| Limited to childhood (n=33) | 23 | 9 | 4 | 0 | 0 | 3 |
| Limited to adulthood (n=13) | 6 | 10 | 0 | 0 | 1 | 0 |

**Supplementary Figure 4. Distribution of modes of inheritances for “Limited to Childhood” and “Limited to Adulthood” conditions.** While autosomal recessive conditions are predominant in “Limited in Childhood” conditions, autosomal dominant conditions are more common in “Limited to Adulthood” conditions.
